# Seroprevalence of IgG antibodies against SARS coronavirus 2 in Belgium – a serial prospective cross-sectional nationwide study of residual samples (March – October 2020)

**DOI:** 10.1101/2020.06.08.20125179

**Authors:** Sereina Herzog, Jessie De Bie, Steven Abrams, Ine Wouters, Esra Ekinci, Lisbeth Patteet, Astrid Coppens, Sandy De Spiegeleer, Philippe Beutels, Pierre Van Damme, Niel Hens, Heidi Theeten

## Abstract

To assess the evolving SARS-CoV-2 seroprevalence and seroincidence related to the national lock-down in Belgium, a nationwide seroprevalence study, stratified by age, sex and region using 3000-4000 residual samples was performed during 7 periods between 30 March and 17 October 2020. Residual sera from ambulatory patients were analyzed for IgG antibodies against S1 proteins of SARS-CoV-2 with a semi-quantitative commercial ELISA. Weighted seroprevalence (overall, by age category and sex) and seroincidence during 7 consecutive periods were estimated for the Belgian population while accommodating test-specific sensitivity and specificity.

The weighted overall seroprevalence initially increased from 1.8% (95% CrI 1.0-2.6) to 5.3% (95% CrI 4.2-6.4), implying a seroincidence of 3.4% (95% CrI 2.4-4.6) between the 1^st^ and 2^nd^ collection period over a period of 3 weeks during the lockdown period (start lockdown mid March 2020). Thereafter, seroprevalence stabilized, however, significant decreases are observed when comparing the 3^rd^ with the 5^th^ and also with the 6^th^ period resulting in negative seroincidence estimates after lockdown was lifted. We estimated for the last collection period mid October 2020 a weighted overall seroprevalence of 4.2% (95% CrI 3.1-5.2).

During lockdown, an initial small but increasing fraction of the Belgian population showed serologically detectable signs of exposure to SARS-CoV-2, which did not further increase when confinement measures eased and full lockdown was lifted.

## Introduction

By August 2021, globally over 213.8 million confirmed cases were reported to be infected by SARS-CoV-2 causing coronavirus disease 2019 (COVID-19). ^1^ Clinical symptoms caused by the virus include loss of taste and smell, fever, malaise, dry cough, shortness of breath, and respiratory distress. Reported illnesses have ranged from very mild to severe (from progressive respiratory failure to death). ^2^ In addition, increasing age, male sex, smoking, and comorbidities such as cardiovascular diseases and diabetes have been identified as risk factors for developing severe illness. ^3^

Until mid-October 2020, the date of the last collection period of this study, there was no vaccine or effective cure available to protect against or treat COVID-19. Therefore, unprecedented measures such as physical distancing, large-scale isolation and closure of borders, schools and workplaces were considered in many countries to mitigate the spread of the disease and to reduce the corresponding pressure on the respective healthcare systems.

In Belgium, the first confirmed COVID-19 case was reported on 4 February 2020, an asymptomatic person repatriated from Wuhan, China. ^4^ The first locally transmitted cases were confirmed on 2 March 2020. Thereafter, the number of confirmed COVID-19 cases rapidly increased. In 2020, Belgium had about 11.49 million inhabitants and is a densely populated European country (374 persons/km^2^) consisting of three regions: Flanders (487 inhabitants/km^2^), Wallonia (216 inhabitants/km^2^), and the Brussels Capital Region (7,501 persons/km^2^).^5^ The Belgian Scientific Institute for Public Health, Sciensano, reported that as of 17 October 2020, 242,217 cases were confirmed (2.1% of the Belgian population; 5.8% of the tested individuals) and 10,410 died. The most affected age category regarding reported cases was 20-29 years (18.3%; 44,411/242,217). ^6^ Importantly, the PCR testing capacity to diagnose SARS-CoV-2 infected subjects in Belgium was very limited during the first weeks of lockdown (2000 – 3000 tests/day) and gradually increased to a more adequate capacity in autumn 2020 (10,000-20,000 tests/day). ^6^ Therefore, more knowledge on and estimation of the age-specific susceptibility to SARS-CoV-2, and its evolution over time, related to control measures that have been taken, is tremendously important to guide policy makers aiming to control the epidemic wave and potential future waves. These needs were translated into the following research objectives: (1) to constitute a national serum bank with residual samples on a periodic basis (cross-sectional study design) in order to estimate the seroprevalence and seroincidence in Belgium and to follow-up trends herein over time and (2) to estimate the age-specific prevalence of SARS-CoV-2 antibodies.

## Methods

### Study design

This prospective cross-sectional nationwide seroprevalence study using residual samples was conducted in individuals aged 0-101 years. In each collection period, sera were collected over one week’s time. The seven collection periods represent different exposure periods: (1) 30 March – 5 April 2020, mainly reflects exposure prior to the lockdown; (2) 20 – 26 April 2020 and (3) 18 – 25 May 2020, additionally reflects exposure during full lockdown; (4) 8 – 13 June 2020, additionally reflects exposure during the period of first relaxation of confinement measures (partial re-opening of schools); (5) 29 June – 4 July 2020, additionally reflects changes during further relaxations (re-opening of shops, restaurants and bars); (6) 7 – 12 September 2020 and (7) 12 – 17 October 2020 are collection periods after the Belgium summer school holidays – see also Figure 1.

**Figure 1.**
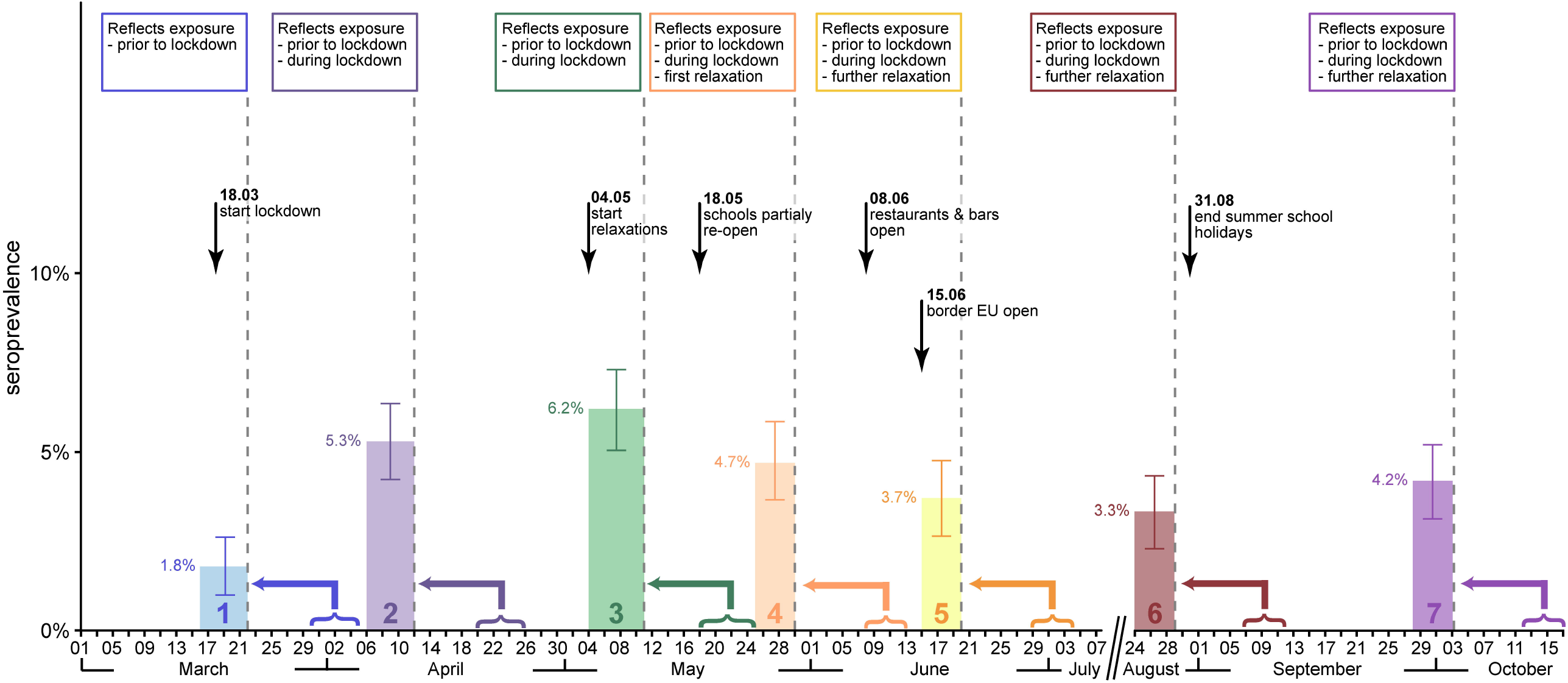
Overview of collection periods related to confinement measures taken in Belgium. Collection periods 1 to 7 are indicated with curly brackets whereas the weighted overall seroprevalence estimates are displayed 14 days earlier in order to reflect the minimum time needed to build up IgG antibodies against SARS-CoV-2 that can be detected by ELISA tests. The bar height shows the weighted overall seroprevalence estimate and the interval the corresponding 95% credible interval.

A serum bank covering all Belgian regions was constituted by collecting residual sera from ten private diagnostic laboratories. The majority of residual samples originate from two large routine laboratories in Flanders (AML) and Wallonia (laboratoire Luc OLIVIER), each with a large geographical network covering the whole of Belgium. Each of these laboratories have a high daily throughput of blood samples, easily receiving up to 23000 samples weekly during the study period for a variety of diagnostics. Each laboratory was allocated a fixed number of samples per age group (10-year age bands, oldest age group ≥90 years), per region (Wallonia, Flanders, Brussels), and per collection period. The number of samples was stratified by sex within each age group.

Residual serum samples in this study originated from ambulatory patients (including people living in nursing homes) visiting their doctor (mainly general practitioners) for any reason including primary care, routine check-up or follow-up of pathology. The samples needed to have a minimum volume of 0.5ml (no hemolysis requirements). To avoid disproportionate selection of subjects with acute and/or severe illness including COVID-19, samples originating from hospitals and triage centers were excluded from the study. Further background information on the residual samples was not available, except for COVID-19 diagnostics. The percentage of COVID-19 diagnostics performed within two weeks prior to collection of the residual samples in this study were calculated in order to check for overrepresentation of residual samples coming from patients who thought they were infected with COVID-19. Only 1-2% of the residual samples collected in period 1 and 2 (PCR test) and 6-8% of the residual samples in period 3-7 (PCR and/or serology test) were linked to COVID-19 diagnostic testing.

### Sample size

The sample size per periodical collection has been calculated according to: (1) previous experience with various age-specific analyses of seroprevalence data in Belgium, ^7^ (2) estimates of the number of COVID-19 infected people in Belgium and (3) the estimated evolution of the epidemic curve. Based on case numbers (hospitalized cases confirmed with COVID-19), the overall prevalence of COVID-19 infection at the start of the study was estimated to be about 0.4%. Therefore, a total sample size of 4,000 in the first collection period ensures the estimation of the overall prevalence with a margin of error of 0.2%; the precision regarding the age-specific prevalence estimates is lower due to the division of samples across the age groups. An increase in prevalence was expected during the study period, as such 3,000 samples from the 2^nd^ collection period onwards were planned. From collection period 2 onwards, target numbers per age group were adapted according to feasibility, sample availability and aiming at maximizing precision and assessing the impact of a change in epidemic control policy.

### Sample preparation and analysis

The mean time between collection and centrifugation of blood samples was 4h, at room temperature. After centrifugation of blood samples, selected residual sera (minimum 0.5 mL) were routinely kept in the fridge (4-8°C) and selected for inclusion and testing within 14 days after collection, according to the manufacturer’s instructions (EuroImmun, Luebeck, Germany), and finally stored at -20°C. The testkit used to obtain serology results was a semi-quantitative test kit (EuroImmun), measuring IgG antibodies against S1 proteins of SARS-CoV-2 in serum (ELISA). The test was performed as previously described by Lassaunière *et al*. ^8^ A case-control validation study with 326 pre-pandemic negative controls and 181 RT-PCR confirmed COVID-19 cases estimated 85.1% sensitivity and 98.8% specificity using the manufacturer’s recommended cutoff for positivity (ratio ≥1.1). ^9^ Presence of detectable IgG antibodies indicates prior exposure to SARS-CoV-2, an infection which may be resolved or is still resolving, and possibly protection against reinfection. ^8,10^

### Data management

Data collected for each sample include: unique sample code, sample date, age, sex, and postal code of the place of residence. Samples were delivered anonymously to the investigators. Triage and check for duplicates was done per collection period in the collecting laboratories before anonymization.

Serological results were linked to the database based on the sample code. No further data entry was required. All files were kept on a secured server at the University of Antwerp. Data will be stored for 20 years.

### Statistical analysis

The serostatus of an individual was considered to be positive if the measured IgG OD values were ≥1.1, equivocal IgG values were considered negative following the manufacturer’s recommendations which were developed for clinical use. Descriptive analysis included mapping of sample origin as well as serostatus (crude figures) up to municipality level per collection period.

For all analyses, the overall seroprevalence, age-specific seroprevalence by 10-year age bands, and seroprevalence by sex for each collection period were obtained as the posterior medians (with 95% credible intervals (CrIs)) of the corresponding posterior distributions for the probabilities to be seropositive. More specifically, a Bayesian approach was considered based on the immunological status (i.e. serostatus) of each individual following a Bernoulli distribution, thereby including individual-specific design weights. Moreover, the model accommodates test-specific sensitivity and specificity of the ELISA assay and uncertainty thereabout when estimating the seroprevalence. In order to inform these quantities, we relied on estimates obtained from the validation study described above. ^9^ The seroincidence estimates were obtained as the posterior medians (with 95% CrIs) of the corresponding posterior distributions for the difference between the probabilities to be seropositive between collection periods. Analytical sensitivity analyses were done: restricting the validation data set to outpatients only, and assuming perfect sensitivity and specificity. The model was implemented in Stan using the interface R (rstan version-2.21.1). ^11,12^ We ran 6000 iterations, convergence was assessed visually and using at the R-hat statistic (more details in Supplementary S1).

We assigned for each collection period weights to the samples such that they mimic the Belgian population structure according to age, sex and provinces for 2020.^5^ Weights are computed by comparing the sample and population frequencies, i.e. we used a complete cross frequency table for sex and 10-year age bands and a marginal distribution for the provinces. Weights were trimmed to a maximum value of 3 to reduce the influence of samples in under-represented strata (Supplementary Figure S1). All analyses were done with the statistical software R (version 4.0.3), to compute weights the package survey (version 4.0) was used. ^13^

### Ethics Committee

The study protocol was approved by the Ethics Committee of the University Hospital Antwerp-University of Antwerp on March 30, 2020 (ref 20/13/158; Belgian Number B3002020000047) and agreed with inclusion without informed consent, on the condition of the samples being collected unlinked and anonymously (see Supplement for study protocol).

## Results

A total of 22,545 serum samples were collected over seven 1 week periods between 30 March and 17 October 2020 to measure the anti-SARS-Cov2 IgG sero-status. The regional, age, and sex distribution of these samples is shown in Table 1 and in Supplementary Figure S2; deviations from the population distribution were taken into account in the estimation of the weighted seroprevalences. Figure 2 shows exemplary for the collection periods 1, 2, and 5 that the origin of the samples was nicely distributed throughout Belgium (panel A-C) and that positive samples were spread over municipalities across Belgium (panel D-F); Supplementary Figures S3-S4 show all collection periods.

**Table 1.** Description of the study population, collection period 1 till 7

|  |  | Collection period 1 |  | Collection period 2 |  | Collection period 3 |  | Collection period 4 |  | Collection period 5 |  | Collection period 6 |  | Collection period 7 |  |
| --- | --- | --- | --- | --- | --- | --- | --- | --- | --- | --- | --- | --- | --- | --- | --- |
|  |  | 30 Mar – 5 Apr 2020 |  | 20 – 26 Apr 2020 |  | 18 – 25 May 2020 |  | 8 – 13 June 2020 |  | 29 June – 4 July 2020 |  | 7 – 12 Sept 2020 |  | 12 – 17 Oct 2020 |  |
|  |  | n | % | n | % | n | % | n | % | n | % | n | % | n | % |
| **Number of samples** |  | 3910 |  | 3397 |  | 3242 |  | 2960 |  | 3023 |  | 3047 |  | 2966 |  |
| **Region** | **Wallonia** | 1511 | 38.6 | 1539 | 45.3 | 1292 | 39.9 | 1100 | 37.2 | 1068 | 35.3 | 1259 | 41.3 | 1144 | 38.6 |
|  | **Flanders** | 2195 | 56.1 | 1556 | 45.8 | 1542 | 47.6 | 1526 | 51.6 | 1621 | 53.6 | 1491 | 48.9 | 1534 | 51.7 |
|  | **Brussels** | 204 | 5.2 | 302 | 8.9 | 408 | 12.6 | 334 | 11.3 | 334 | 11.0 | 297 | 9.7 | 288 | 9.7 |
| **Age in years** | **0-10** | 36 | 0.9 | 85 | 2.5 | 174 | 5.4 | 124 | 4.2 | 110 | 3.6 | 68 | 2.2 | 68 | 2.3 |
|  | **10-20** | 294 | 7.5 | 442 | 13.0 | 431 | 13.3 | 375 | 12.7 | 413 | 13.7 | 432 | 14.2 | 405 | 13.7 |
|  | **20-30** | 436 | 11.2 | 375 | 11.0 | 414 | 12.8 | 383 | 12.9 | 394 | 13.0 | 406 | 13.3 | 402 | 13.6 |
|  | **30-40** | 461 | 11.8 | 407 | 12.0 | 424 | 13.1 | 395 | 13.3 | 396 | 13.1 | 396 | 13.0 | 397 | 13.4 |
|  | **40-50** | 468 | 12.0 | 406 | 12.0 | 411 | 12.7 | 394 | 13.3 | 403 | 13.3 | 399 | 13.1 | 397 | 13.4 |
|  | **50-60** | 498 | 12.7 | 430 | 12.7 | 419 | 12.9 | 393 | 13.3 | 400 | 13.2 | 402 | 13.2 | 400 | 13.5 |
|  | **60-70** | 507 | 13.0 | 426 | 12.5 | 417 | 12.9 | 399 | 13.5 | 403 | 13.3 | 403 | 13.2 | 406 | 13.7 |
|  | **70-80** | 506 | 12.9 | 316 | 9.3 | 236 | 7.3 | 201 | 6.8 | 204 | 6.7 | 212 | 7.0 | 204 | 6.9 |
|  | **80-90** | 493 | 12.6 | 315 | 9.3 | 163 | 5.0 | 166 | 5.6 | 160 | 5.3 | 167 | 5.5 | 160 | 5.4 |
|  | **≥90** | 211 | 5.4 | 195 | 5.7 | 153 | 4.7 | 130 | 4.4 | 140 | 4.6 | 162 | 5.3 | 127 | 4.3 |
| **Sex** | **male** | 1799 | 46.0 | 1599 | 47.1 | 1587 | 49.0 | 1425 | 48.1 | 1471 | 48.7 | 1500 | 49.2 | 1377 | 46.4 |
|  | **female** | 2111 | 54.0 | 1798 | 52.9 | 1655 | 52.0 | 1535 | 51.9 | 1552 | 51.3 | 1547 | 50.8 | 1589 | 53.6 |

**Figure 2.**
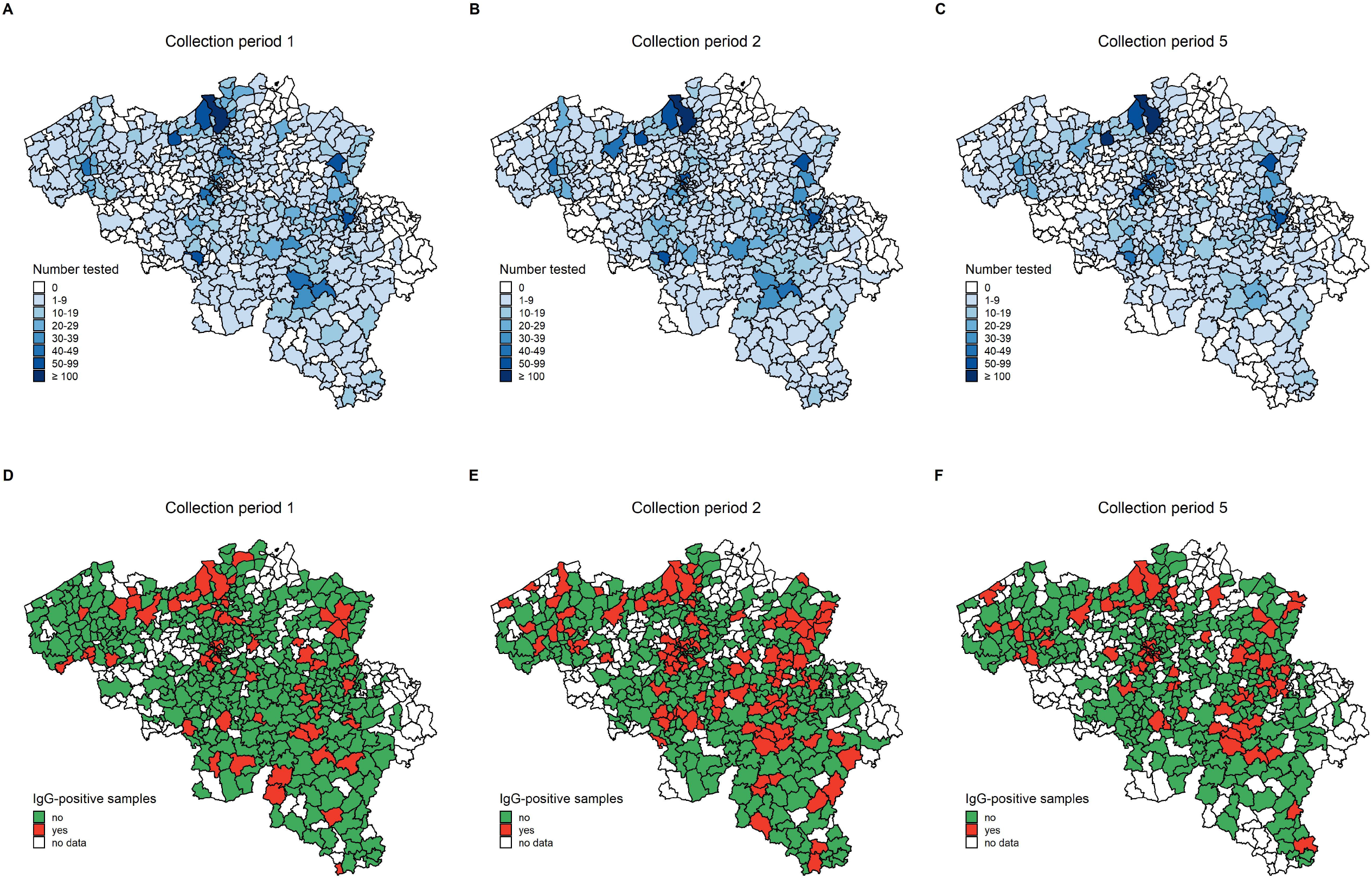
Map of Belgium at municipality level for collection period 1, 2, and 5. Panel A-C: number of samples tested in each municipality; panel D-F: presence of IgG-positive (red) versus exclusively IgG-negative (green) samples in each municipality.

At the start, the seroprevalence estimates per age category ranged between 0.6% (20-30 years) and 5.9% (0-10 years) in collection period 1. The weighted overall seroprevalence showed a significant increase between collection period 1 and 2, i.e. from 1.8% (95% CrI 1.0-2.6) to 5.3% (95% CrI 4.2-6.4) over a period of 3 weeks (Figure 3, panel A) which is also shown by the overall seroincidence estimate of 3.4% (95% CrI 2.4-4.6) (Figure 3, panel D). This significant increase in seroprevalence is reflected in the age categories 20-49, 80-89, and ≥90 as indicated by the seroincidence estimates (Figure 3, panel B+E) and within each sex (Figure 3, panel C+F).

**Figure 3.**
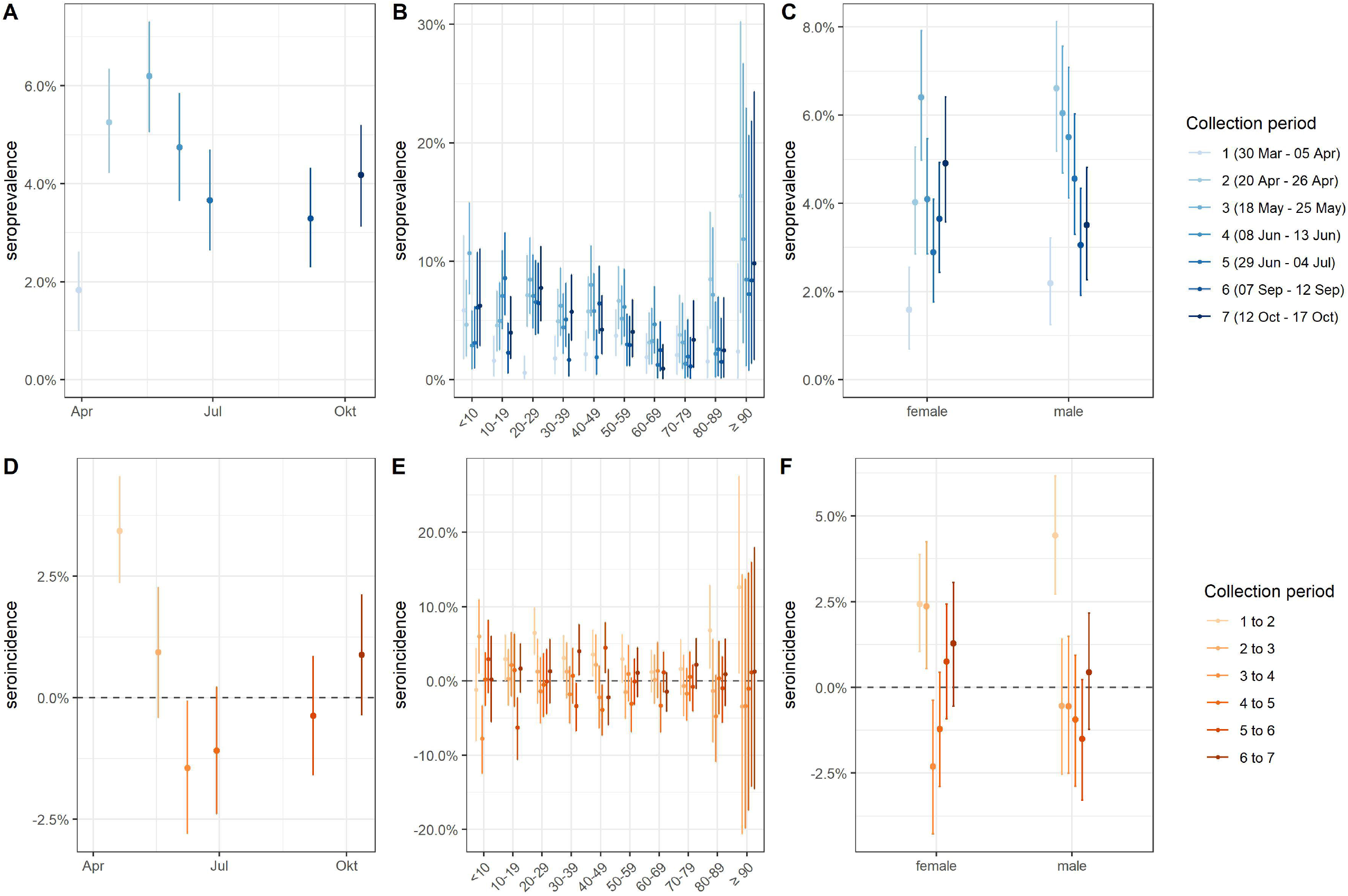
Weighted seroprevalence (A, B, C) and seroincidence (D, E, F) estimates in Belgium. Overall (panel A+D), by 10-year age bands (panel B+E), by sex (panel C+F): estimates (point) with the corresponding 95% credible intervals (lines). Note, the seroincidence estimates indicate the difference in seroprevalence estimates between two consecutive collection rounds.

In comparison with period 2, the overall seroprevalence stabilized thereafter until period 4 as shown in Figure 3 (panel A). For the last collection period mid-October an overall seroprevalence of 4.2% (95% CrI 3.1-5.2) was estimated. However, significant decreases are observed when comparing the 3^rd^ period with the 5^th^, 6^th^ and 7^th^: seroincidence of -2.5% (95% CrI -1.2 to -3.9), -2.9% (95% CrI -1.6 to -4.2), and -2.0% (95% CrI -0.7 to -3.4). For the first two comparisons, these decreases were also observed for three subgroups: age categories <10, 40-49, and females. Comparing males and females, we observed significantly higher seroprevalence estimate in period 2 for males (+2.6%; 95% CrI 0.7-4.5), and from 2^nd^ to 3^rd^ period a higher seroincidence estimate for females (+2.9%; 95% CrI 1.1-4.5). For these analyses, the diagnostic test-specific sensitivity and specificity were estimated to be 84.9% (95% CrI 82.9-86.9) and 98.7% (95% CrI 98.2-99.1), respectively. ^9^ The analytical sensitivity analysis results, which investigated the influence of sensitivity and specificity, i.e. once restricting the validation data set to outpatients only, and once assuming perfect sensitivity and specificity, do not change the interpretation (Supplementary Figures S5-S10).

## Discussion

This study estimates seroprevalence and seroincidence of IgG antibodies against S1 proteins of SARS-CoV-2 in Belgium based on a total of 22,545 residual sera collected in seven rounds from 30 March – 17 October 2020. The results give an indication of the state of the COVID-19 epidemic in Belgium, showing that only an estimated 1.8% (95% CrI 1.0-2.6) of the population had detectable antibodies against SARS-CoV-2 at the start of lockdown (March 2020), which had more than doubled to 5.3% (95% CrI 4.2-6.4) three weeks later (seroincidence +3.4%, 95% CrI 2.4-4.6). However, seroprevalence stabilized thereafter and decreased until start of summer holidays (July) to 3.7% (95% CrI 2.6-4.7), which was also reflected by negative seroincidence estimates. These seroprevalences continued in the same range, even after re-opening of the schools after summer resulting in a seroprevalence of 4.2% (95% CrI 3.1-5.2) by mid-October.

Stringent containment measures were enforced in Belgium as of 13 March 2020. These included travel bans, closures of borders, schools, shops, factories and social gatherings in an effort to contain the spread of COVID-19 and decrease the pressure on health care systems. These intervention measures slowed down the number of COVID-19 patients that were hospitalized daily. In the first two weeks of the lockdown (up to 25 March 2020), over 500 cases were hospitalized daily, and this growth rate halved four weeks later. ^6^ By 26 April 2020, 0.1% of the Belgian population had been hospitalized for COVID-19 (14,822/11.46×10^6^) and 0.4% of the Belgian population had tested positive for SARS-CoV-2 (48,093/11.46×10^6^) on a total of 32,1862 screened patients. ^6^ The estimated seroprevalence (5.3%, 95% CrI 4.2-6.4) in the same period (20-26 April 2020) indicates that far more people had generated antibodies against SARS-CoV-2 and thus had been in contact with the virus than what was expected from the number of COVID-19 confirmed cases reported in Belgium at that time. These seroprevalences provide insights into the dark number of SARS-CoV-2 infections, which is indeed a multiple of the confirmed cases. By end of June, the number of daily hospital admissions in Belgium dropped below 20 and the number of confirmed COVID-19 cases stabilized at a lower level than the estimated seroprevalence in Belgium but hospital admission increased again by October (Table 2). Clearly, the reported numbers of COVID-19 confirmed cases represent an underestimation and were influenced by the testing policy as testing was initially focused on the most severe symptomatic cases, presenting to hospitals. Vice versa, also the seroprevalences in this study are possibly underestimated because residual samples from hospitals were excluded. Moreover, patients with upper respiratory tract infections were not allowed to visit general practitioners and ambulatory care during the lockdown period, possibly contributing to further underestimation of the seroprevalences in collection periods 2, 3 and 4. The risk population, who possibly adhered better to self-confinement, as well as patients with non-urgent health problems were less likely to visit their doctor until later stages of the epidemic (collection periods 5-7). This may result in a higher proportion of non-COVID-19 infected subjects of whom residual samples have been analyzed in the later collection periods, hence contributing, at least partly, to a significant drop in seroprevalences and seroincidence towards the 5^th^ collection period (see also Stringhini et al., 2020^14^). Regardless of this change in care seeking behavior throughout the study, the current seroprevalence study in combination with the reported confirmed COVID-19 cases may form a useful tool to estimate the total number of recently acquired SARS-CoV-2 infections in Belgium. Moreover, this study gives an indication of the seroprevalence and seroincidence in light of the confinement measures taken in Belgium which helps understanding the spread of SARS-CoV-2 and the significance of periodical variations.

**Table 2.** Number of confirmed COVID-19 cases versus weighted seroprevalence in Belgium during the different collection periods

|  | Collection period | Confirmed hospitalized cases per day<sup>s</sup> | Testing strategy for COVID-19<sup>§</sup> | Total confirmed COVID-19 cases* | Weighted seroprevalence (with 95% CrI) | Weighted seroincidence<sup>#</sup> (with 95% CrI) |
| --- | --- | --- | --- | --- | --- | --- |
| 1 | 30 Mar – 5 Apr 2020 | 510.3 | Only asymptomatic cases and health care workers (~38400 weekly new tests; 28.2% positivity rate) | 23252/11.46 milj = 0.2% | 1.8% (1.0 to 2.6) | - |
| 2 | 20 – 26 Apr 2020 | 202.9 | Including National testing platform in elderly homes (~106000 weekly new tests; 7.1% positivity rate) | 48093/11.46 milj = 0.4% | 5.3% (4.2 to 6.4) | 3.4% (2.4 to 4.6) |
| 3 | 18 – 25 May 2020 | 51.8 | Including testing of all suspected COVID-19 cases (~86000 weekly new tests; 2.6% positivity rate) | 58134/11.46 milj = 0.5% | 6.2% (5.1 to 7.3) | 0.9% (-0.4 to 2.3) |
| 4 | 8 – 13 Jun 2020 | 21.8 | (~87000 weekly new tests; 1.2% positivity rate) | 60460/11.46 milj = 0.5% | 4.7% (3.7 to 5.9) | -1.4% (-2.8 to -0.1) |
| 5 | 29 Jun – 4 Jul 2020 | 12.5 | (~85000 weekly new tests; 0.9% positivity rate) | 62306/11.46 milj = 0.5% | 3.7% (2.6 to 4.7) | -1.1% (-2.4 to 0.2) |
| 6 | 9 – 12 Sept 2020 | 31.2 | (~214000 weekly new tests; 3.1% positivity rate) | 94908/11.46 milj = 0.8% | 3.3% (2.3 to 4.3) | -0.4% (-1.6 to 0.9) |
| 7 | 12 – 17 Oct 2020 | 263.0 | (~419000 weekly new tests; 16.3% positivity rate) | 242217/11.46 milj = 2.1% | 4.2% (3.1 to 5.2) | 0.9% (-0.4 to 2.1) |

<sup>s</sup>average over the collection period, data reported by Sciensano<sup>6</sup>; <sup>§</sup>weekly new tests and positivity rate reported for the same week number by Sciensano<sup>6</sup>; \*reported
at last day of collection period by Sciensano<sup>6</sup>; <sup>#</sup>in comparison with previous collection period

From the above it is clear that determining of the extent of spread of SARS-CoV-2 at country level is a challenge. Moreover, the sensitivity of the serological test used in this study depends on the time since the onset of symptoms, ^8,15^ thereby preventing a fraction of the infected subjects to test seropositive if not infected long enough or too long prior to testing. By day 14 after symptom onset, IgG against SARS-CoV-2 are detectable in serum of the majority of patients. ^2^ Possibly, recent SARS-CoV-2 infected subjects may have been included in the current study of whom antibodies were not yet detectable in blood. Asymptomatic and pauci-symptomatic SARS-CoV-2 infected subjects of whom it is reported that they may develop low or no antibodies against SARS-CoV-2, may have been included in this study as well. ^16^ Some studies reported that the decay of IgG levels starts within 2-3 months^17–19^ but others reported antibody response to be long lasting. ^20,21^ If IgG levels really decrease within few months this could result in the situation that both pauci-symptomatic subjects as well as subjects that suffered from an infection more than a few months ago may have received a seronegative test result at the time point of collection and thus cause underestimation of incidence of infection. Moreover, this would also imply that seroprevalence studies on SARS-CoV-2 would only be able to give information on the past few months.

A seroprevalence study conducted in healthy adult blood donors in Belgium described, similar to this study, a doubling of seroprevalence estimates between end of March and mid-April, which was followed by stable estimates around 5% by mid-September. ^22^ In Switzerland, a population-based household study conducted in Geneva also estimated also a doubling of seroprevalence estimates within five weeks (6 April – 9 May 2020) from 4·8% (95% CI 2.4-8.0, n=341) to 10·8% (95% CI 8.2-13.9, n=775). ^14^ In Spain, a national seroprevalence of 5% (95% CI 4.7-5.4, n=61,075) was reported in the period from 27 April – 11 May 2020, i.e. comparable magnitude as in our study. ^23^ In the UK, the plateauing of seroprevalences has also been observed in a study with blood donors in June 2020.^24^ However, comparing seroprevalence estimates across countries are hampered due to several reasons, for example, differences in policy and intervention measures taken initially, the extent to which social activity levels and the social contact behaviour in general have changed due to stringent lockdown measures, the relaxations thereof over time, study designs, etc. A dashboard called SeroTracker has been developed in order to easily visualize global SARS-CoV-2 seroprevalence estimates. ^25^ They report over 2000 seroprevalence estimates from 112 countries, indicating that our study is one of the few that reports several rounds of seroprevalence estimates at a national level for the general population.

## Conclusion

Serial seroprevalence monitoring indicates that in Belgium, a densely populated country in the center of Western Europe, SARS-COV-2 virus was introduced all over the country from the start and the proportion of the infected seropositive population at least doubled within 3 weeks’ time from 1.8% to 5.3% during the start of the lockdown in spring 2020. In line with reported confirmed cases and COVID-19 deaths, estimated seroprevalence plateaued and seroincidence decreased thereafter. The observed decay of the proportion seropositives by the end of June would corroborate with reports of quick antibody waning after mild or asymptomatic infection^17-19^ but could also be an issue of change in care seeking behavior. Serial seroprevalence and seroincidence monitoring in combination with COVID-19 diagnostic testing data can provide a useful tool to estimate the proportion of the population recently infected with SARS-CoV-2. These findings may have helped to calibrate the Belgian response to the epidemic’s first wave and to guide policy makers to control for potential future waves.

### Data sharing

Data is available on Zenodo (http://doi.org/10.5281/zenodo.4664403).

## Supporting information

STROBE checklist

## Data Availability

Data is available on Zenodo (http://doi.org/10.5281/zenodo.4664403).

## Acknowledgments

We acknowledge the Belgian laboratories that voluntarily collected sera and data for this study: Algemeen Medisch Laboratorium (AML, Antwerpen), Laboratoire Luc OLIVIER (Fernelmont), Declerck Klinisch Laboratorium (Ardooie), Klinisch Labo RIGO (Genk), Labo Anacura/Nuytinck (Evergem), Labo Somedi (Heist-op-den-Berg), Labo LBS (Brussels), Laboratoire Bauduin (Enghien), Medisch labo Bruyland (Kortrijk), Synlab (Luik).

