## Supplemental material for "Seroprevalence of IgG antibodies against SARS coronavirus 2 in Belgium – a serial prospective cross-sectional nationwide study of residual samples (March – October 2020)"

| | | |
|---|---|---|
| **Study Protocol Sero-Epidemiology** | | 3 |
| **S1.** | Details on statistical framework | 11 |
| **Figure S1** | Distribution of weights by collection period | 13 |
| **Figure S2.** | Age distribution of Belgium population and samples by sex | 14 |
| **Figure S3.** | Map of Belgium at municipality level, collection period 1 till 4 | 15 |
| **Figure S4.** | Map of Belgium at municipality level, collection period 5 till 7 | 16 |
| **Table S1.** | Values of weighted seroprevalence estimates displayed in Figure 3 (main text) | 17 |
| **Table S2.** | Values of weighted seroincidence estimates displayed in Figure 3 (main text) | 19 |
| **Figure S5.** | Analytical sensitivity analysis - weighted seroprevalence estimates in Belgium overall | 21 |
| **Figure S6.** | Analytical sensitivity analysis - weighted seroprevalence estimates in Belgium by 10-year age bands | 22 |
| **Figure S7.** | Analytical sensitivity analysis - weighted seroprevalence estimates in Belgium by sex | 23 |
| **Figure S8.** | Analytical sensitivity analysis - weighted seroincidence estimates in Belgium overall | 24 |
| **Figure S9.** | Analytical sensitivity analysis - weighted seroincidence estimates in Belgium by 10-year age bands | 25 |
| **Figure S10.** | Analytical sensitivity analysis - weighted seroincidence estimates in Belgium by sex | 26 |

This supplementary material has been provided by the authors to give readers additional information about their work.

**Study Protocol Sero-Epidemiology.**

Study Protocol sero-epidemiology COVID-19 Belgium 2020

1. GENERAL INFORMATION

The study will be coordinated by the Centre for the Evaluation of Vaccination, Vaccine & Infectious Disease Institute, at the University of Antwerp (UAntwerpen) (Heidi Theeten, Pierre Van Damme). Sample collection will be coordinated by AML-RIATOL, in collaboration with laboratoire Olivier.

Partners:
Serum samples will be collected by voluntarily participating laboratories; i.c AML (Algemeen Medisch Laboratorium; Emiel Vloorsstraat 9, 2020 Antwerpen) and Olivier Laboratories ( Rue Léopold Genicot 16, 5380 Fernelmont).
CHERMID, at the University of Antwerp (UAntwerpen) (Philippe Beutels, Sereina Herzog) and UHasselt (Niel Hens, Steven Abrams, ) provide methodological support.

2. OBJECTIVE OF THE STUDY

#### 2.1 Description of overall objective

The overall objective of the study is to constitute a national bank of sera on a periodic basis (every 3 weeks from April 2020 onward) in order to estimate the age-specific seroprevalence of emerging COVID-19 biomarkers in Belgium and to follow-up trends therein.

At first, the study aims to estimate the age-specific prevalence of SARS-CoV-2 antibodies in Belgium, in order to identify groups that have been infected versus those at risk of infection as a function of time. These analyses are planned in 2020, which is also the year of collection of the serum bank.

3. SCIENTIFIC RELEVANCE

#### 3.1 Scientific background

The COVID-19 pandemic started in Wuhan, China, with several patients (cluster of size 41) with pneumonia, developing symptoms between December 8, 2019 and January 2, 2020, which all visited a local market. COVID-19 disease is caused by a novel coronavirus (severe acute respiratory syndrome-related coronavirus 2, or SARS-CoV-2) likely originating from bats due to its genetic similarity to bat coronaviruses. Coronaviruses are a large family of viruses that are common in humans and many different species, other family members are the Middle East respiratory syndrome-related coronavirus (MERS-CoV) and the severe acute respiratory syndrome-related coronavirus (SARS-CoV).

After cases were reported in Wuhan, the disease started spreading throughout the world, being officially called a pandemic by WHO on March 11, 2020.
The virus causing COVID-19 is easily spreading from person to person. Cases as well as community spread is being detected in most countries worldwide as there is no pre-existing immunity against this new virus.

This is the first known pandemic outbreak caused by the emergence of a new coronavirus. Pandemics of respiratory diseases follow a defined evolution ('Pandemic Intervals Framework'). It starts with an investigation phase followed by the recognition, the initiation and the acceleration phase. The end of the acceleration phase is determined by the peak of illnesses and is followed by the deceleration phase (implying a decrease in illnesses). Different countries as well as different parts of the same country can be in different phases of a pandemic.

The risk of infection depends on the characteristics of the virus (including the spreadability of the virus), the severity of resulting illness and the medical availability to control the relative success of these factors.

#### 3.2 Public health relevance

The expansive transmission of COVID-19 could translate into a large number of medical care at the same time with elevated hospitalization and deaths. Public health and healthcare systems may become overloaded and healthcare providers and hospitals may be overwhelmed. At his moment, there is no vaccine to protect against COVID-19 disease and no medications approved to treat COVID-19.

The complete clinical outcome with regarding COVID-19 is not fully known. Reported illnesses have ranged from very mild to severe, including illness resulting in death. In addition, older people and people of all ages with severe chronic medical conditions, like heart disease, lung disease and diabetes, seem to be at higher risk of developing severe COVID-19 illness.

The first cases of COVID-19 infection in Belgium that were not related to Chinese cases, were reported in the first week of March 2020. The virus is extremely contagious, but transmission dynamics are largely unknown. Probably children are involved in transmission, but severe cases in children are rarely reported. Non-pharmaceutical interventions become the most important strategy to try to delay the spread and the impact of diseases.
Control measures considered to prevent mitigation are, amongst others, closing schools and daycare centers for children. However, the economic and educational impact of these measures is considerable, and their effectiveness to prevent or delay transmission remains to date unknown.

In order to guide policy to control the current outbreak and eventual future relapses, knowledge of the age-specific susceptibility to SARS-CoV-2 and its evolution over time as related to control measures that have been taken is tremendously important. The current study will document the increasing background immunity in the population by age-group, and the remaining vulnerable proportion in the population over time.

### 4. DEFINITIONS AND ABBREVIATIONS

Immunity will be expressed as the proportion of each age group having a SARS-CoV-2-antibody concentration above the cut-off described by the manufacturer of the assay.

### 5. METHODS

#### Overall study design: description of the overall study and methods

A serum bank will be constituted based on a prospective cross-sectional study design. Sera will be obtained by using the residual sera collected during routine laboratory testing in collaborating laboratories.
Serum specimens submitted to diagnostic laboratories may not be entirely representative of the population, but they are readily accessible and therefore feasible to collect.

#### 5.1 Study design

The study population is the population living in Belgium, with a serum sample analysed at one of the participating laboratories during the study period.
To avoid overselection of subjects with acute and/or severe illness, samples collected in hospitals will not be selected. During the CoVid19 containment period (from 13/3/2020 onward), health care in hospitals is reduced and all non-urgent care is being delayed or

referred to outpatient services. Moreover, data on CoVid19 in hospitalised patients are available as far as actual screening guidelines were applicable for them.

#### 5.2 Sampling

To cover all regions of Belgium, two large private diagnostic laboratories were approached to collect serum samples, one for the Flemish part and Brussels and one for the French speaking part. Each laboratory will be allocated a fixed number of samples per age group and per periodical collection period. The number of samples will further be stratified by gender, to obtain equal numbers of males and females in each age group. Each periodical collection will last 1 week. This way of sampling avoids working with hospitals or general practitioners (sentinel), and will not cause additional workload for hospitals and general practices.

The sample size per periodical collection has been calculated according to previous experience with various age-specific analyses of seroprevalence data in Belgium, and according to estimates of the number of CoVid19 infected people in Belgium and of the estimated evolution of the epidemic curve. Based on case numbers (hospitalized cases confirmed with COVID-19), the overall prevalence of COVID-19 infection at start of the study is estimated to be about 0.4%. A total sample size of 4000 allows an overall estimate with a margin of error of 0.2%; precision of age-specific prevalence estimates will be lower. However, an increase in prevalence is expected during the study period. If the overall prevalence increases to 4% in all age groups, a sample number of 400 per age category allows a margin of error of 2%.

Age groups will be defined in 10 year age bands (0-9; 10-19; …). In total, up to 14000 sera will be collected, distributed over the periodical collections as presented in Table 1.

Table 1: Number of samples per age group

| Collection period (in 2020) | Relative number | Total number |
|---|---|---|
| 1 (30/3-5/4) | 400 per age group | 4000 |
| 2 (20/4-26/4) | 300 per age group | 3000 |
| 3 (11/5-17/5) | 300 per age group | 3000 |
| 4 (1/6-7/6) | 200 per age group | 2000 |
| 5 (22/6-28/6) | 200 per age group | 2000 |

If these numbers would not be feasible, either for collecting or for testing, an alternative strategy is to focus on selected age categories instead of the whole age range, i.e. 0-5; 10-15; 20-25; 35-40; 60+. Prevalence in the in-between age categories can be extrapolated based on the available data in their neighboring age categories, and on the age-specific proportion of confirmed COVID-19 cases (case-based reporting to health authorities). Such strategy increases efficiency, especially in a low-prevalence situation.

#### 5.3 Data management

*Data collection and data source*
Samples will be delivered **unlinked and anonymous** to the investigators. Triage and check for duplicates will be done in the collecting laboratories. Samples will be assigned a unique study code number (different from the laboratory code) which can be linked only to the limited set of background data that is extracted for the study. For each sample, only the unique sample code, age (in years), sample date, gender, postal code of the place of residence, and sample type (serum/plasma/other) will be provided to the CEV by the laboratory. By doing so

the researchers will not be able to identify the patient or owner of the sample. The region (Brussels, Flanders, Wallonia) and province of residence will be derived from the postal code. No specific training on data collection is required. Collecting laboratories will receive one pre-labeled tube per sample to include in the study.

*Data flow and management*

Every collection period, the information collected on the samples by the two laboratories will be sent to the CEV by email, through an excel sheet (Annex 1). Data will be checked for completeness (based on age, gender and postal code) by a staff member of the CEV. When needed, laboratories will be contacted to collect a new sample within the same collection period.

Separate files per laboratory will be held until completion of the sample collection. They will be merged into one database once the results for serological analysis are available. Serology results (SARS-CoV2 antibodies) will be linked to the database based on the sample code. No further data entry is required. All files will be kept on a secured server at Uantwerpen, with restricted access. Data will be stored for 20 years

*Data analysis*

To estimate overall prevalence rates, the prevalence of seronegativity, seropositivity and equivocal results will be standardized for age and gender according to the Belgian population structure, based on the most recent available National Registry data and 95% confidence intervals will be calculated per age group.

Logistic regression will evaluate the effect of age, gender and region upon the serostatus.

#### 5.4 Test objects and samples

The samples collected will be serum . An amount of 2 ml (minimum 0.5 ml) is requested.

Processing of blood samples after sampling will be according to local standards of the laboratories, but residual serum/plasma selected for the collection should be stored in the fridge (4-8°C) after centrifugation for a maximum of 4 days and then kept at -20° at the collecting laboratory until transport to the analysing laboratory. Samples will be kept at -20° (on dry ice) during transport.

Samples in which results are equivocal will be retested once before being assigned to the protected or susceptible or equivocal category as specified by the manufacturer of the test used.

After analysis, samples will be stored ultimately until end of 2020.

### 6. Scientific Review

A scientific steering committee will be set up to assist in follow-up of the progress of the study, and to validate and discuss results.

### 7. Organisation of the research project

#### 7.1 Starting and completion date

The collection of serum samples will start at approval until the required number of samples is achieved.

#### 7.2 Subcontracting

No subcontracting is initiated so far. The collaborating laboratories will select, store and deliver samples pro bono, at this stage, as no budget is yet foreseen for this project.

### 8. Resources

No funding so far

### 9. Risk and benefits for participants and informed consent requirements

The study protocol is submitted for approval to the Ethical committee of the University of Antwerp . Since data are provided **unlinked anonymous** and blood **samples are taken for diagnostic purposes without any extra amount of blood being drawn**, an informed consent of the persons sampled to take part in this study is not requested.
Individual coordinates are not collected, therefore participating individuals cannot be informed about the results of the serological test (unlinked anonymous). Sample donors will thus not have any benefit nor risk from the fact that their sample is included.

### 10. Property rights of study material and results

The results of the study are the property of the CEV. Transmission of the results to external institutions is submitted to internal rules of the CEV.

**Annex 1: Data collection sheet for laboratories**

Laboratory Nb 1

| Code | Sample date | Age (Years) | Gender | Postal code | Sample type |
|---|---|---|---|---|---|
| 01/F/0/01 | | | | | |
| 01/F/0/02 | | | | | |
| 01/F/0/03 | | | | | |
| 01/M/0/01 | | | | | |
| …. | | | | | |

### S1. Details on statistical framework

We want to estimate the seroprevalence for each collection period and the corresponding seroincidence between the collection periods: overall, stratified by 10-year age bands, and stratified by sex.

The Bayesian model mentioned in the main text considers the immunological status (i.e. serostatus) of each individual, which follows a Bernoulli distribution, thereby including binomial models for the IgG ELISA assay specific sensitivity and specificity. More specifically, the probability of observing a positive test result is assumed to be a function of the true positive rate (i.e., sensitivity of the diagnostic test), the false positive rate (i.e., (1 – specificity) of the test) and the true underlying probability of seropositivity for an individual. This model structure presumes an invariant sensitivity and specificity of the diagnostic test, irrespective of the timing of the test (relative to symptom onset or infection) and sample collection. In addition, we use individual-specific design weights in the log-likelihood function such that for each collection period the observations mimic the Belgian population structure according to age, sex and provinces for 2020 (see main text for a detailed description).

$$y_{i,g} \sim Bernoulli\left(p_g * SE + \left(1 - p_g\right) * (1 - SP)\right)$$
$$x^+ \sim Binomial(n^+, SE)$$
$$x^- \sim Binomial(n^-, 1 - SP)$$

$$\mathcal{LL} = \sum_{g=1}^{G} \sum_{i=1}^{I_g} \omega\left(c_{i,g}, a_{i,g}, s_{i,g}, l_{i,g}\right) * \left[y_{i,g}\, log\left(p_g^*\right) + \left(1 - y_{i,g}\right) * log\left(1 - p_g^*\right)\right] + x^+ log(SE) + (n^+ - x^+) * log(1 - SE) + x^- log(1 - SP) + (n^- - x^-) * log(SP)$$

with

- $\boldsymbol{y_{i,g}}$ being the serostatus for the i[th] observation in group $g$,
- $\boldsymbol{p_g}$ being the probability to be seropositive in group $g$,
- $\boldsymbol{SE}$ and $\boldsymbol{SP}$ being the IgG ELISA assay specific sensitivity and specificity,
- $\boldsymbol{p_g^*} = p_g * SE + \left(1 - p_g\right) * (1 - SP)$,
- $\boldsymbol{\omega}\left(c_{i,g}, a_{i,g}, s_{i,g}, p_{i,g}\right)$ being the individual-specific design weight depending on the collection period $c$, age category $a$, sex $s$, and province $l$ observation $i$ of group $g$ belongs to,
- $\boldsymbol{G}$ being the number of groups $g$ investigated in the analysis,
- $\boldsymbol{I_g}$ being the number of observations which belong to the same group $g$,

Observations in group $g$ consists of:

- *overall*: all observations who belong to the same collection period,
- *by age*: all observations who belong to the same collection period and age category,
- *by sex*: all observations who belong to the same collection period and sex.

We assumed that the sensitivity ($SE$) and specificity ($SP$) are the same for all observations. The sensitivity, $SE$, is determined using $n^+$ positive controls from the case-control validation study by Meyer *et al.*, of which $x^+$ tested positive. The specificity, $SP$, is determined using $n^+$ pre-pandemic negative controls, of which $x^-$ tested positive, see *Table S1.1*.[1] As mentioned above, this model structure presumes an invariant sensitivity and specificity of the diagnostic test, irrespective of the timing of the test (relative to symptom onset or infection) and sample collection

We used U(0, 1)-priors (uniform priors on the unit interval) for the sensitivity and specificity of the diagnostic test and a Beta(1,1)-priors for the probabilities to be seropositive. We implemented this model in the Stan probabilistic programming language using the interface R, i.e. R package rstan (version 2.21.1).[2] We ran 6000 iterations (6 chains with 1500 iterations each with 500 for warm-up). Convergence was assessed visually and using R-hat statistic.

We used for the main analysis in our study all positive controls (n=181) and for the technical sensitivity analysis only the outpatients (n=90) and once a model without considering sensitivity and specificity. *Table S1.2* shows the model estimates for the diagnostic test-specific sensitivity and specificity.

We computed for each iteration the difference between two consecutive seroprevalence estimates in order to get seroincidence estimates between each collection period.

For all analyses, the overall seroprevalence, age-specific seroprevalence by 10-year age bands, and seroprevalence by sex for each collection period were obtained as the posterior medians (with 95% credible intervals (CrIs)) of the corresponding posterior distributions for the probabilities to be seropositive. The seroincidence estimates were obtained as the posterior medians (with 95% CrIs) of the corresponding posterior distributions for the difference between the probabilities to be seropositive between the collection periods.

***Table S1.1.*** Data used to determine sensitivity and specificity from case-control validation study Meyer *et al.*[1]

| Samples | Total | ELISA IgG EI cut-offs[§] | | |
|---|---|---|---|---|
| | | Negative | Indeterminate | Positive |
| Negative controls | 326 | 314 | 8 | 4 |
| Positive controls | | | | |
| COVID-19 - all patient | 181 | 26 | 1 | 154 |
| COVID-19 – only outpatients | 90 | 14 | 0 | 76 |

[§]EI cut-offs: <0.8 = negative; ≥ 0.8 and < 1.1 = indeterminate; ≥ 1.1 = positive

***Table S1.2.*** The median estimated sensitivity and specificity of the model for the main and analytical sensitivity analyses based on the case-control validation data.

| Analysis | Positive controls | | Negative controls | | Sensitivity (95% CrI) | Specificity (95% CrI) |
|---|---|---|---|---|---|---|
| | Total | Tested positive | Total | Tested positive | | |
| Main | 181 | 154 | 326 | 4 | 84.9% (82.9 – 86.9) | 98.7% (98.2 – 99.1) |
| Analytical sensitivity | 90 | 76 | 326 | 4 | 84.2% (81.2 – 87.0) | 98.8% (98.3 – 99.1) |

CrI: credible interval, i.e. the 2.5th and 97.5th percentiles of the posterior distribution

**Figure S1.** Distribution of weights by collection period.

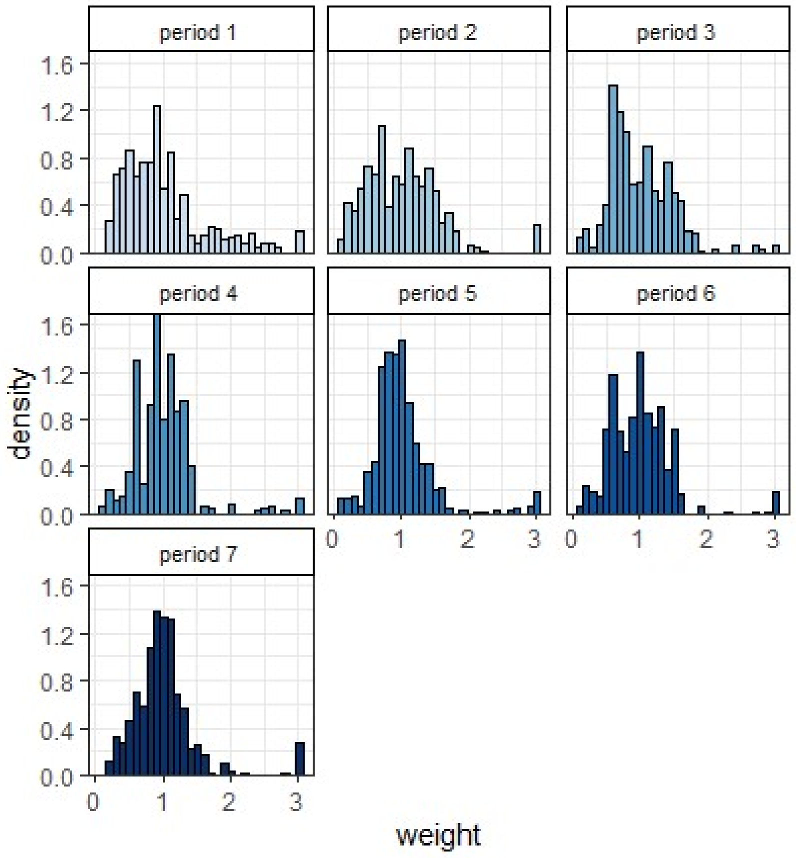

**Figure S2.** Age distribution of Belgium population (population) and samples (period 1 till 7) by sex. Bars within each panel sum up to 100%.

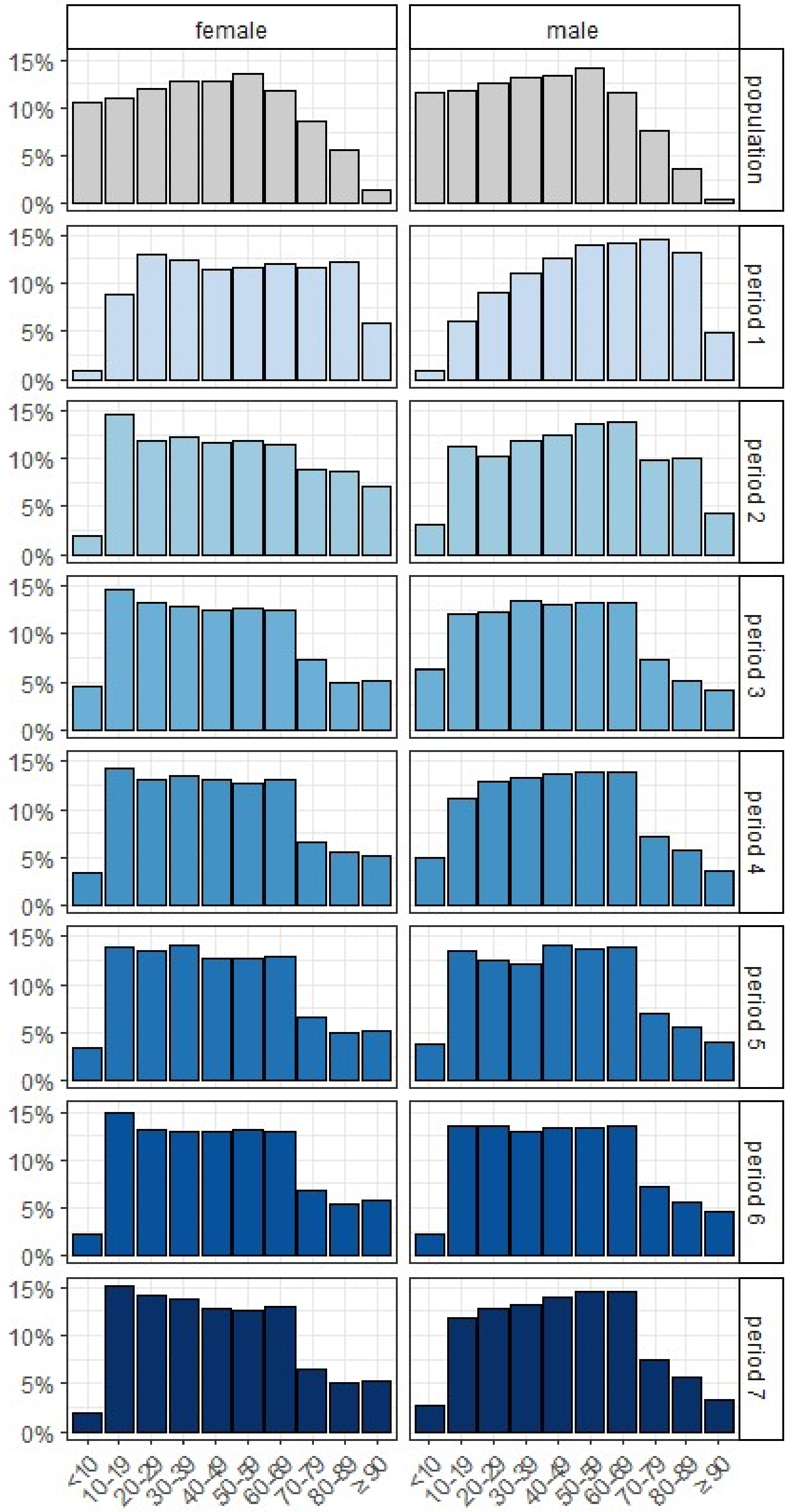

**Figure S3.** Map of Belgium at municipality level, collection period 1 till 4; panel A-D: number of samples tested in each municipality, panel E-H: presence of IgG-positive (red) versus exclusively IgG-negative (green) samples in each municipality.

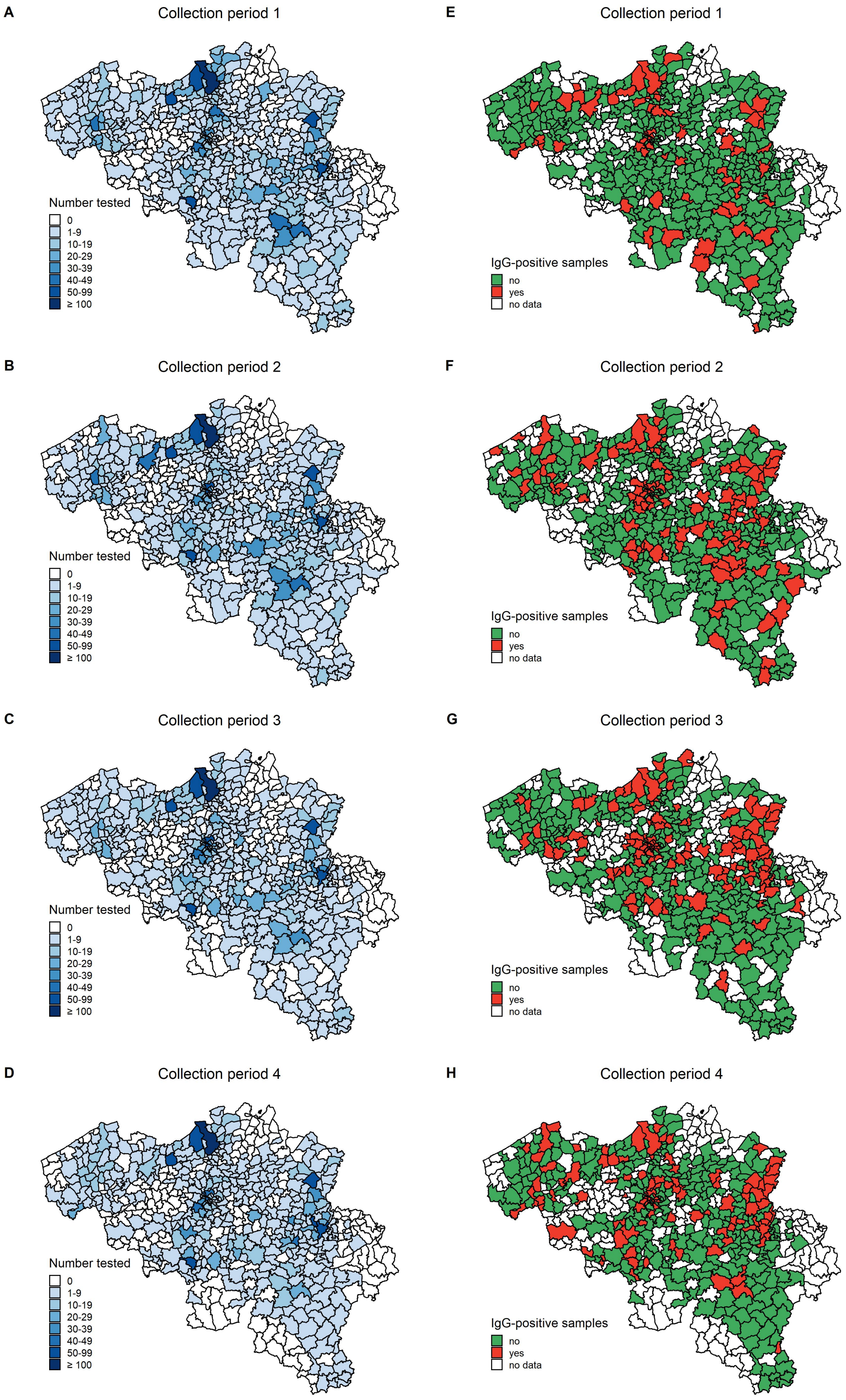

**Figure S4.** Map of Belgium at municipality level, collection period 5 till 7; panel A-C: number of samples tested in each municipality, panel D-F: presence of IgG-positive (red) versus exclusively IgG-negative (green) samples in each municipality.

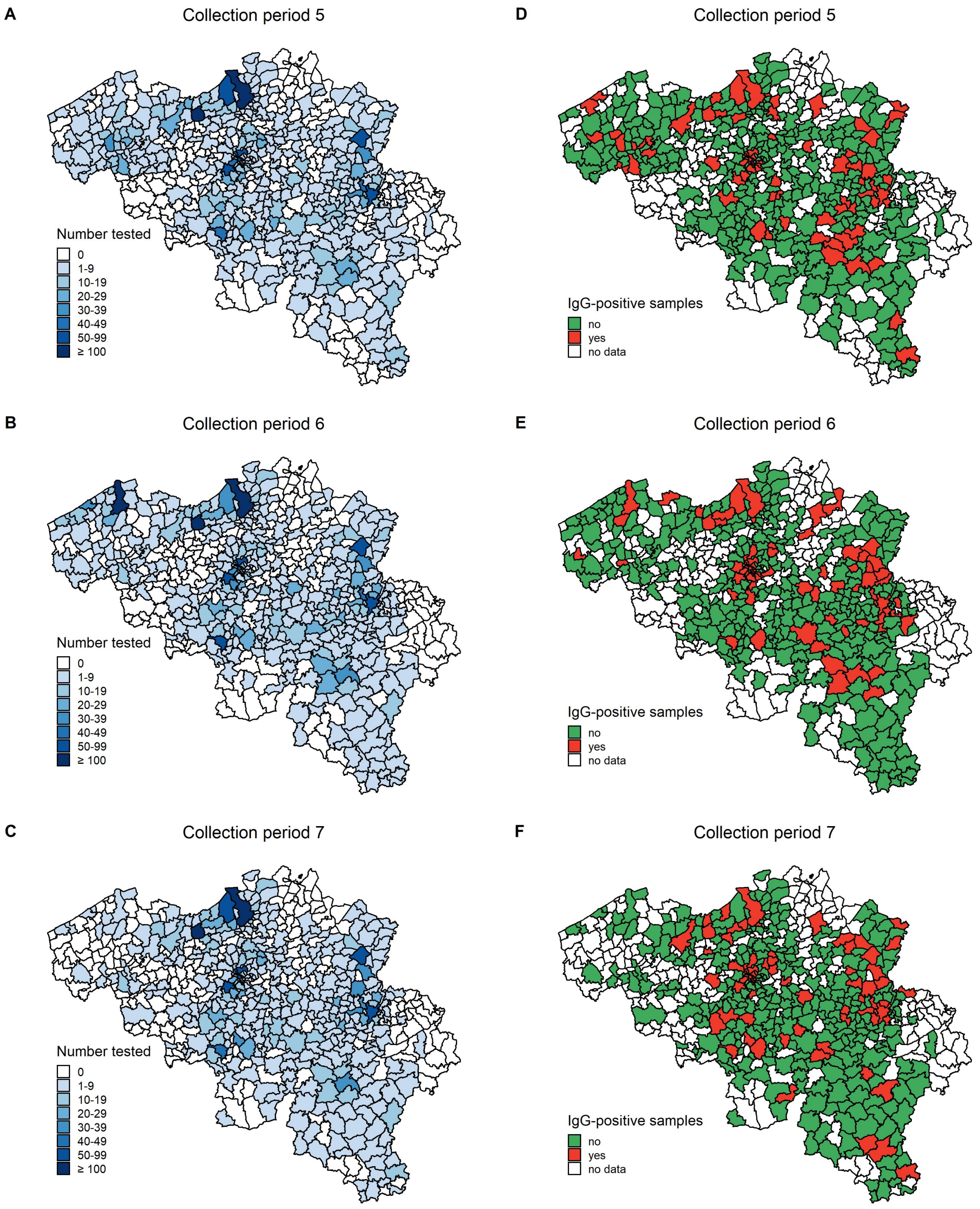

**Table S1.** Weighted seroprevalence in Belgium overall, by 10-year age bands, and by sex as displayed in Figure 3 (main text).

| collection period | analysis | level | median | 2.5% | 97.5% |
|---|---|---|---|---|---|
| 1 | overall | - | 1.83 | 1.00 | 2.61 |
| 2 | overall | - | 5.25 | 4.22 | 6.35 |
| 3 | overall | - | 6.20 | 5.05 | 7.31 |
| 4 | overall | - | 4.74 | 3.65 | 5.85 |
| 5 | overall | - | 3.66 | 2.64 | 4.70 |
| 6 | overall | - | 3.28 | 2.29 | 4.32 |
| 7 | overall | - | 4.18 | 3.12 | 5.20 |
| 1 | age | [0,10) | 5.86 | 1.75 | 12.15 |
| 1 | age | [10,20) | 1.62 | 0.25 | 3.70 |
| 1 | age | [20,30) | 0.58 | 0.03 | 2.00 |
| 1 | age | [30,40) | 1.80 | 0.45 | 3.69 |
| 1 | age | [40,50) | 2.16 | 0.72 | 4.12 |
| 1 | age | [50,60) | 3.72 | 2.00 | 5.94 |
| 1 | age | [60,70) | 1.91 | 0.48 | 3.91 |
| 1 | age | [70,80) | 2.09 | 0.43 | 4.55 |
| 1 | age | [80,90) | 1.53 | 0.12 | 4.47 |
| 1 | age | 90+ | 2.38 | 0.09 | 9.78 |
| 2 | age | [0,10) | 4.66 | 1.95 | 8.41 |
| 2 | age | [10,20) | 4.59 | 2.37 | 7.48 |
| 2 | age | [20,30) | 7.14 | 4.45 | 10.49 |
| 2 | age | [30,40) | 4.95 | 2.79 | 7.62 |
| 2 | age | [40,50) | 5.78 | 3.49 | 8.68 |
| 2 | age | [50,60) | 6.67 | 4.27 | 9.57 |
| 2 | age | [60,70) | 3.13 | 1.27 | 5.66 |
| 2 | age | [70,80) | 3.79 | 1.41 | 7.15 |
| 2 | age | [80,90) | 8.47 | 4.29 | 14.16 |
| 2 | age | 90+ | 15.52 | 5.64 | 30.24 |
| 3 | age | [0,10) | 10.70 | 7.22 | 14.93 |
| 3 | age | [10,20) | 4.98 | 2.51 | 8.18 |
| 3 | age | [20,30) | 8.44 | 5.53 | 11.99 |
| 3 | age | [30,40) | 6.26 | 3.71 | 9.43 |
| 3 | age | [40,50) | 8.01 | 5.34 | 11.31 |
| 3 | age | [50,60) | 5.15 | 2.96 | 7.95 |
| 3 | age | [60,70) | 3.29 | 1.29 | 6.05 |
| 3 | age | [70,80) | 3.13 | 0.90 | 6.46 |
| 3 | age | [80,90) | 7.17 | 3.11 | 12.81 |
| 3 | age | 90+ | 11.87 | 3.07 | 26.70 |
| 4 | age | [0,10) | 2.89 | 0.88 | 5.82 |
| 4 | age | [10,20) | 7.09 | 4.27 | 10.91 |
| 4 | age | [20,30) | 7.07 | 4.31 | 10.57 |
| 4 | age | [30,40) | 4.41 | 2.20 | 7.26 |
| 4 | age | [40,50) | 5.81 | 3.30 | 8.96 |
| 4 | age | [50,60) | 6.15 | 3.66 | 9.33 |
| 4 | age | [60,70) | 4.68 | 2.22 | 7.86 |
| 4 | age | [70,80) | 1.36 | 0.10 | 4.18 |
| 4 | age | [80,90) | 2.20 | 0.19 | 6.58 |
| 4 | age | 90+ | 8.44 | 1.13 | 22.92 |
| 5 | age | [0,10) | 3.11 | 0.98 | 6.16 |
| 5 | age | [10,20) | 8.58 | 5.44 | 12.41 |
| 5 | age | [20,30) | 6.56 | 3.77 | 10.07 |
| 5 | age | [30,40) | 5.09 | 2.77 | 8.17 |
| 5 | age | [40,50) | 1.89 | 0.43 | 4.21 |
| 5 | age | [50,60) | 3.00 | 1.17 | 5.54 |
| 5 | age | [60,70) | 1.24 | 0.11 | 3.43 |
| 5 | age | [70,80) | 1.96 | 0.23 | 5.05 |
| 5 | age | [80,90) | 2.58 | 0.31 | 6.99 |
| 5 | age | 90+ | 7.23 | 0.78 | 20.59 |
| 6 | age | [0,10) | 6.08 | 2.68 | 10.76 |
| 6 | age | [10,20) | 2.29 | 0.52 | 4.82 |
| 6 | age | [20,30) | 6.46 | 3.87 | 9.86 |
| 6 | age | [30,40) | 1.66 | 0.28 | 3.87 |
| 6 | age | [40,50) | 6.44 | 3.91 | 9.59 |
| 6 | age | [50,60) | 2.93 | 1.16 | 5.34 |
| 6 | age | [60,70) | 2.49 | 0.71 | 4.91 |
| 6 | age | [70,80) | 1.12 | 0.06 | 3.60 |
| 6 | age | [80,90) | 1.50 | 0.09 | 5.19 |
| 6 | age | 90+ | 8.40 | 1.34 | 21.81 |

| collection period | analysis | level | median | 2.5% | 97.5% |
|---|---|---|---|---|---|
| 7 | age | [0,10) | 6.25 | 2.85 | 11.03 |
| 7 | age | [10,20) | 3.96 | 1.77 | 7.01 |
| 7 | age | [20,30) | 7.76 | 4.93 | 11.27 |
| 7 | age | [30,40) | 5.74 | 3.29 | 8.88 |
| 7 | age | [40,50) | 4.22 | 2.15 | 7.11 |
| 7 | age | [50,60) | 4.04 | 1.89 | 6.76 |
| 7 | age | [60,70) | 0.95 | 0.06 | 2.99 |
| 7 | age | [70,80) | 3.36 | 1.01 | 6.70 |
| 7 | age | [80,90) | 2.46 | 0.21 | 6.91 |
| 7 | age | 90+ | 9.82 | 1.66 | 24.30 |
| 1 | sex | male | 2.19 | 1.25 | 3.22 |
| 1 | sex | female | 1.59 | 0.69 | 2.54 |
| 2 | sex | male | 6.61 | 5.18 | 8.13 |
| 2 | sex | female | 4.02 | 2.86 | 5.28 |
| 3 | sex | male | 6.05 | 4.68 | 7.57 |
| 3 | sex | female | 6.41 | 4.98 | 7.91 |
| 4 | sex | male | 5.51 | 4.12 | 7.08 |
| 4 | sex | female | 4.09 | 2.86 | 5.46 |
| 5 | sex | male | 4.56 | 3.30 | 6.03 |
| 5 | sex | female | 2.89 | 1.76 | 4.10 |
| 6 | sex | male | 3.05 | 1.91 | 4.34 |
| 6 | sex | female | 3.65 | 2.44 | 4.93 |
| 7 | sex | male | 3.51 | 2.26 | 4.82 |
| 7 | sex | female | 4.91 | 3.57 | 6.42 |

Note, age category “[a,b)” means that “a ≤ age < b”.

**Table S2.** Weighted seroincidence in Belgium overall, by 10-year age bands, and by sex as displayed in Figure 3 (main text).

| comparing periods | analysis | level | median | 2.5% | 97.5% |
|---|---|---|---|---|---|
| 1 to 2 | overall | - | 3.43 | 2.36 | 4.56 |
| 2 to 3 | overall | - | 0.93 | -0.42 | 2.28 |
| 3 to 4 | overall | - | -1.44 | -2.80 | -0.06 |
| 4 to 5 | overall | - | -1.09 | -2.39 | 0.22 |
| 5 to 6 | overall | - | -0.37 | -1.60 | 0.86 |
| 6 to 7 | overall | - | 0.88 | -0.36 | 2.12 |
| 1 to 2 | age | [0,10) | -1.17 | -8.12 | 4.35 |
| 1 to 2 | age | [10,20) | 2.94 | -0.03 | 6.23 |
| 1 to 2 | age | [20,30) | 6.46 | 3.56 | 9.81 |
| 1 to 2 | age | [30,40) | 3.12 | 0.18 | 6.17 |
| 1 to 2 | age | [40,50) | 3.58 | 0.61 | 6.89 |
| 1 to 2 | age | [50,60) | 2.93 | -0.25 | 6.28 |
| 1 to 2 | age | [60,70) | 1.21 | -1.52 | 4.11 |
| 1 to 2 | age | [70,80) | 1.65 | -1.83 | 5.55 |
| 1 to 2 | age | [80,90) | 6.81 | 1.66 | 12.87 |
| 1 to 2 | age | 90+ | 12.63 | 1.00 | 27.58 |
| 2 to 3 | age | [0,10) | 6.02 | 0.99 | 10.95 |
| 2 to 3 | age | [10,20) | 0.32 | -3.28 | 4.25 |
| 2 to 3 | age | [20,30) | 1.29 | -3.01 | 5.61 |
| 2 to 3 | age | [30,40) | 1.29 | -2.35 | 5.16 |
| 2 to 3 | age | [40,50) | 2.19 | -1.68 | 6.28 |
| 2 to 3 | age | [50,60) | -1.49 | -5.08 | 2.06 |
| 2 to 3 | age | [60,70) | 0.14 | -3.06 | 3.48 |
| 2 to 3 | age | [70,80) | -0.67 | -4.67 | 3.46 |
| 2 to 3 | age | [80,90) | -1.34 | -8.28 | 5.59 |
| 2 to 3 | age | 90+ | -3.40 | -20.61 | 14.30 |
| 3 to 4 | age | [0,10) | -7.74 | -12.48 | -3.33 |
| 3 to 4 | age | [10,20) | 2.11 | -2.02 | 6.51 |
| 3 to 4 | age | [20,30) | -1.39 | -5.71 | 3.13 |
| 3 to 4 | age | [30,40) | -1.82 | -5.71 | 1.91 |
| 3 to 4 | age | [40,50) | -2.18 | -6.37 | 2.02 |
| 3 to 4 | age | [50,60) | 0.97 | -2.78 | 4.86 |
| 3 to 4 | age | [60,70) | 1.36 | -2.31 | 5.18 |
| 3 to 4 | age | [70,80) | -1.67 | -5.35 | 1.70 |
| 3 to 4 | age | [80,90) | -4.75 | -10.90 | 0.82 |
| 3 to 4 | age | 90+ | -3.37 | -19.83 | 13.70 |
| 4 to 5 | age | [0,10) | 0.21 | -3.30 | 3.81 |
| 4 to 5 | age | [10,20) | 1.48 | -3.49 | 6.33 |
| 4 to 5 | age | [20,30) | -0.50 | -4.86 | 3.73 |
| 4 to 5 | age | [30,40) | 0.71 | -3.04 | 4.41 |
| 4 to 5 | age | [40,50) | -3.87 | -7.36 | -0.50 |
| 4 to 5 | age | [50,60) | -3.08 | -6.84 | 0.39 |
| 4 to 5 | age | [60,70) | -3.34 | -6.91 | -0.19 |
| 4 to 5 | age | [70,80) | 0.56 | -2.73 | 3.92 |
| 4 to 5 | age | [80,90) | 0.34 | -4.48 | 5.32 |
| 4 to 5 | age | 90+ | -1.05 | -17.46 | 14.55 |
| 5 to 6 | age | [0,10) | 2.92 | -1.57 | 8.12 |
| 5 to 6 | age | [10,20) | -6.27 | -10.58 | -2.27 |
| 5 to 6 | age | [20,30) | -0.07 | -4.42 | 4.29 |
| 5 to 6 | age | [30,40) | -3.39 | -6.77 | -0.26 |
| 5 to 6 | age | [40,50) | 4.47 | 1.08 | 7.90 |
| 5 to 6 | age | [50,60) | -0.06 | -3.15 | 2.93 |
| 5 to 6 | age | [60,70) | 1.18 | -1.53 | 3.91 |
| 5 to 6 | age | [70,80) | -0.76 | -4.08 | 2.17 |
| 5 to 6 | age | [80,90) | -0.96 | -5.62 | 3.27 |
| 5 to 6 | age | 90+ | 1.17 | -14.23 | 15.97 |
| 6 to 7 | age | [0,10) | 0.17 | -5.49 | 6.02 |
| 6 to 7 | age | [10,20) | 1.67 | -1.61 | 5.04 |
| 6 to 7 | age | [20,30) | 1.32 | -3.02 | 5.59 |
| 6 to 7 | age | [30,40) | 4.04 | 0.77 | 7.57 |
| 6 to 7 | age | [40,50) | -2.21 | -5.94 | 1.54 |
| 6 to 7 | age | [50,60) | 1.12 | -2.16 | 4.47 |
| 6 to 7 | age | [60,70) | -1.44 | -4.14 | 1.14 |
| 6 to 7 | age | [70,80) | 2.16 | -1.08 | 5.75 |
| 6 to 7 | age | [80,90) | 0.89 | -3.38 | 5.66 |
| 6 to 7 | age | 90+ | 1.26 | -14.53 | 17.92 |
| 1 to 2 | sex | male | 4.42 | 2.72 | 6.17 |

| comparing periods | analysis | level | median | 2.5% | 97.5% |
|---|---|---|---|---|---|
| 1 to 2 | sex | female | 2.43 | 1.05 | 3.87 |
| 2 to 3 | sex | male | -0.54 | -2.53 | 1.40 |
| 2 to 3 | sex | female | 2.36 | 0.55 | 4.23 |
| 3 to 4 | sex | male | -0.55 | -2.50 | 1.49 |
| 3 to 4 | sex | female | -2.31 | -4.28 | -0.37 |
| 4 to 5 | sex | male | -0.93 | -2.88 | 0.93 |
| 4 to 5 | sex | female | -1.21 | -2.89 | 0.44 |
| 5 to 6 | sex | male | -1.50 | -3.29 | 0.22 |
| 5 to 6 | sex | female | 0.75 | -0.92 | 2.43 |
| 6 to 7 | sex | male | 0.44 | -1.23 | 2.16 |
| 6 to 7 | sex | female | 1.28 | -0.55 | 3.06 |

Note, age category “[a,b)” means that “a ≤ age < b”.

**Figure S5.** Analytical sensitivity analysis - weighted seroprevalence estimates in Belgium overall

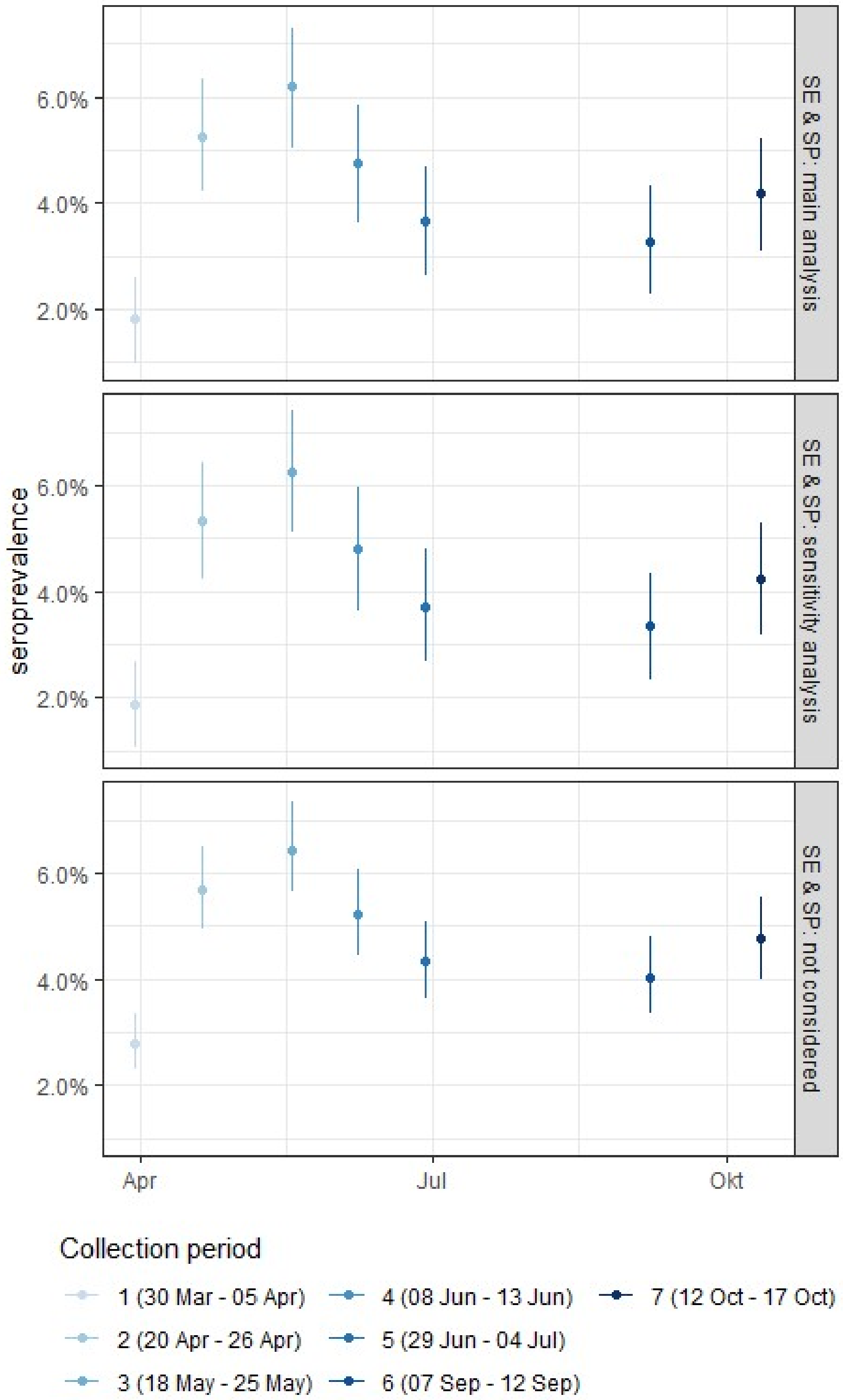

**Panel 'SE & SP: main analysis'** contains the estimates displayed in the main text (Figure 3)
**Panel 'SE & SP: sensitivity analysis'** contains the analytical sensitivity results which uses only information given for outpatient positive controls (n=90) for estimating sensitivity and specificity
**Panel 'SE & SP: not considered'** contains the estimates if sensitivity and specificity are not considered in the analysis

**Figure S6.** Analytical sensitivity analysis - weighted seroprevalence estimates in Belgium by 10-year age bands

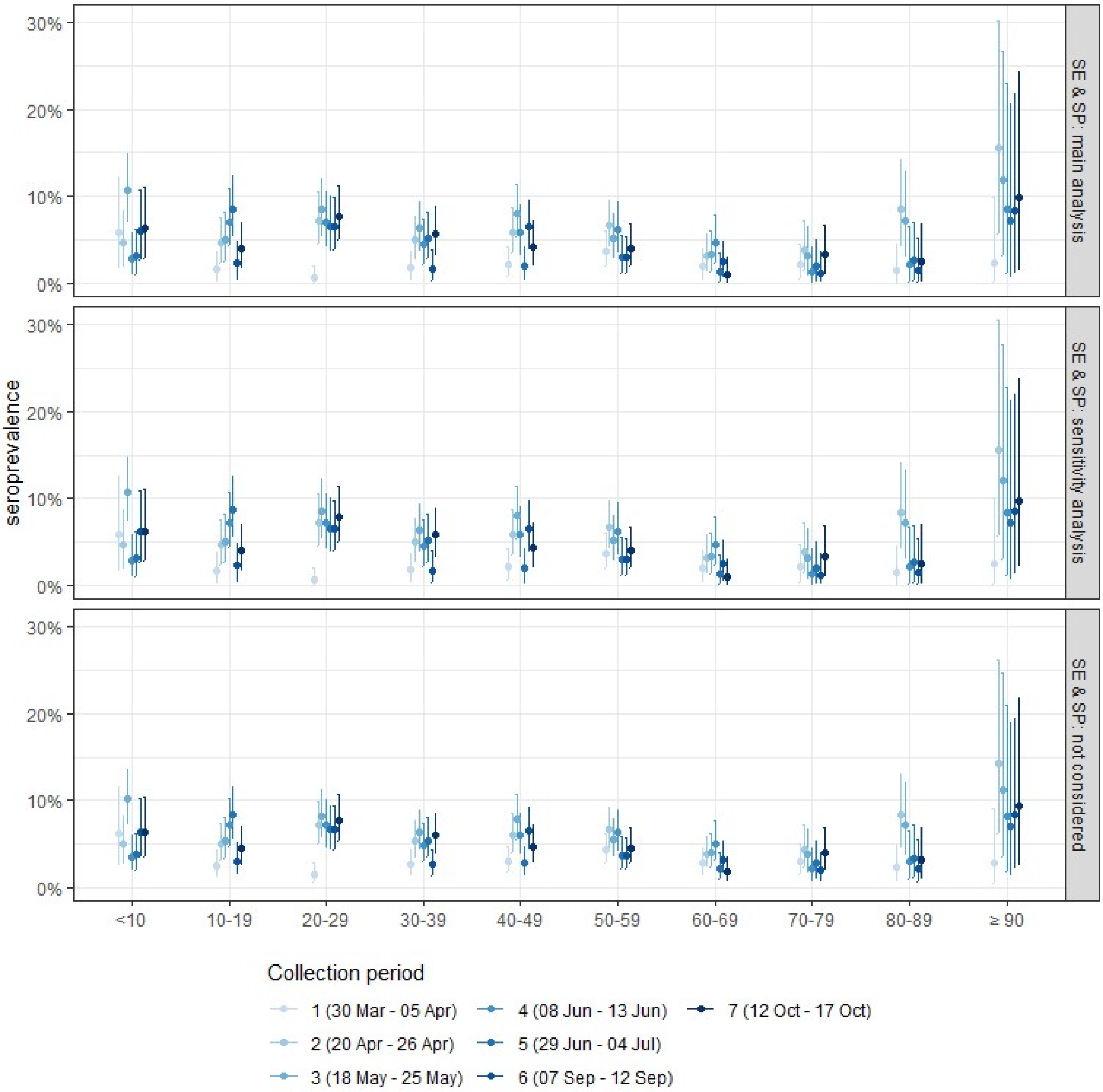

**Panel 'SE & SP: main analysis'** contains the estimates displayed in the main text (Figure 3)
**Panel 'SE & SP: sensitivity analysis'** contains the analytical sensitivity results which uses only information given for outpatient positive controls (n=90) for estimating sensitivity and specificity
**Panel 'SE & SP: not considered'** contains the estimates if sensitivity and specificity are not considered in the analysis

**Figure S7.** Analytical sensitivity analysis - weighted seroprevalence estimates in Belgium by sex

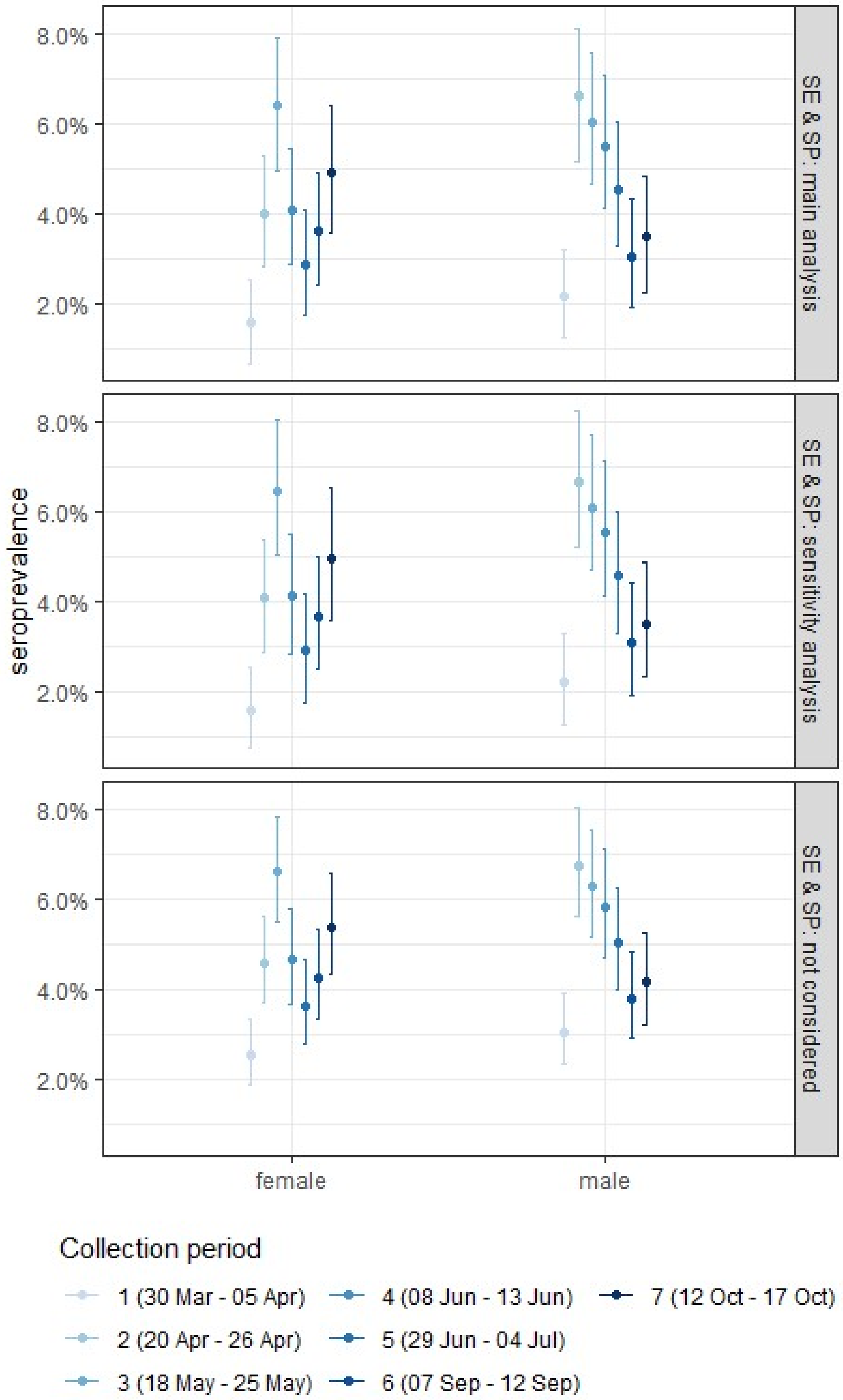

**Panel 'SE & SP: main analysis'** contains the estimates displayed in the main text (Figure 3)
**Panel 'SE & SP: sensitivity analysis'** contains the analytical sensitivity results which uses only information given for outpatient positive controls (n=90) for estimating sensitivity and specificity
**Panel 'SE & SP: not considered'** contains the estimates if sensitivity and specificity are not considered in the analysis

**Figure S8.** Analytical sensitivity analysis - weighted seroincidence estimates in Belgium overall

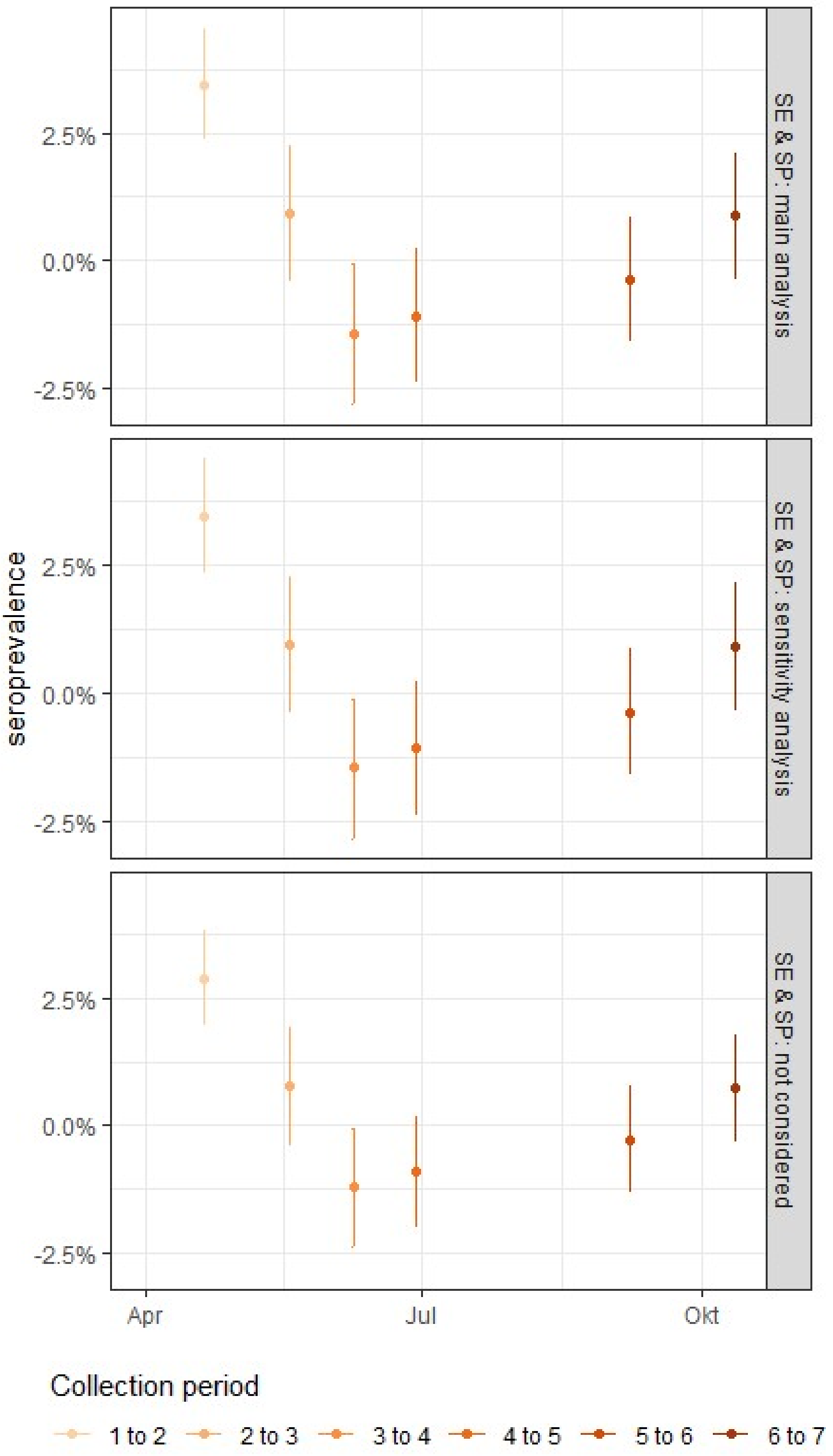

**Panel 'SE & SP: main analysis'** contains the estimates displayed in the main text (Figure 3)
**Panel 'SE & SP: sensitivity analysis'** contains the analytical sensitivity results which uses only information given for outpatient positive controls (n=90) for estimating sensitivity and specificity
**Panel 'SE & SP: not considered'** contains the estimates if sensitivity and specificity are not considered in the analysis

**Figure S9.** Analytical sensitivity analysis - weighted seroincidence estimates in Belgium by 10-year age bands

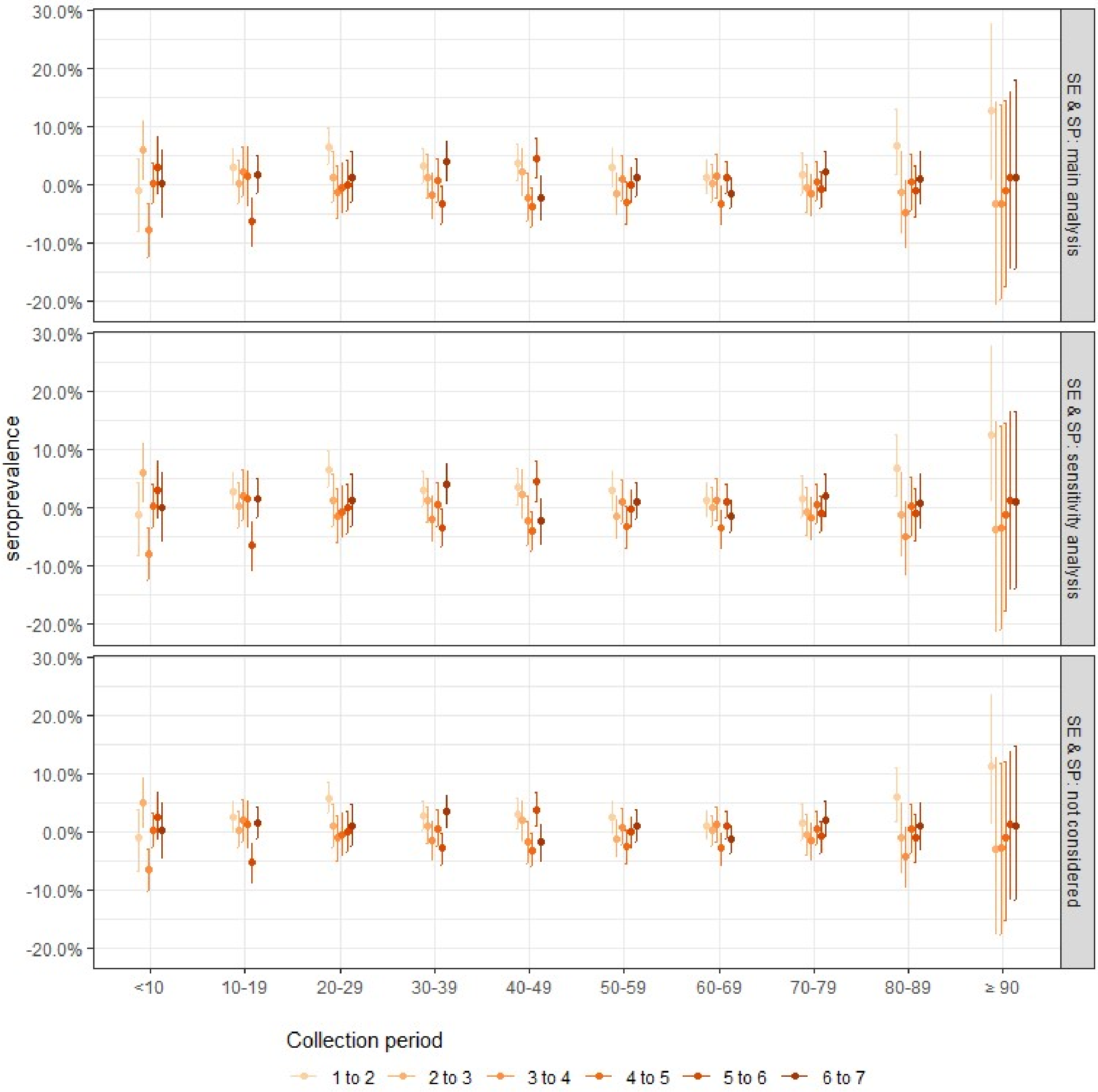

**Panel 'SE & SP: main analysis'** contains the estimates displayed in the main text (Figure 3)
**Panel 'SE & SP: sensitivity analysis'** contains the analytical sensitivity results which uses only information given for outpatient positive controls (n=90) for estimating sensitivity and specificity
**Panel 'SE & SP: not considered'** contains the estimates if sensitivity and specificity are not considered in the analysis

**Figure S10.** Analytical sensitivity analysis - weighted seroincidence estimates in Belgium by sex

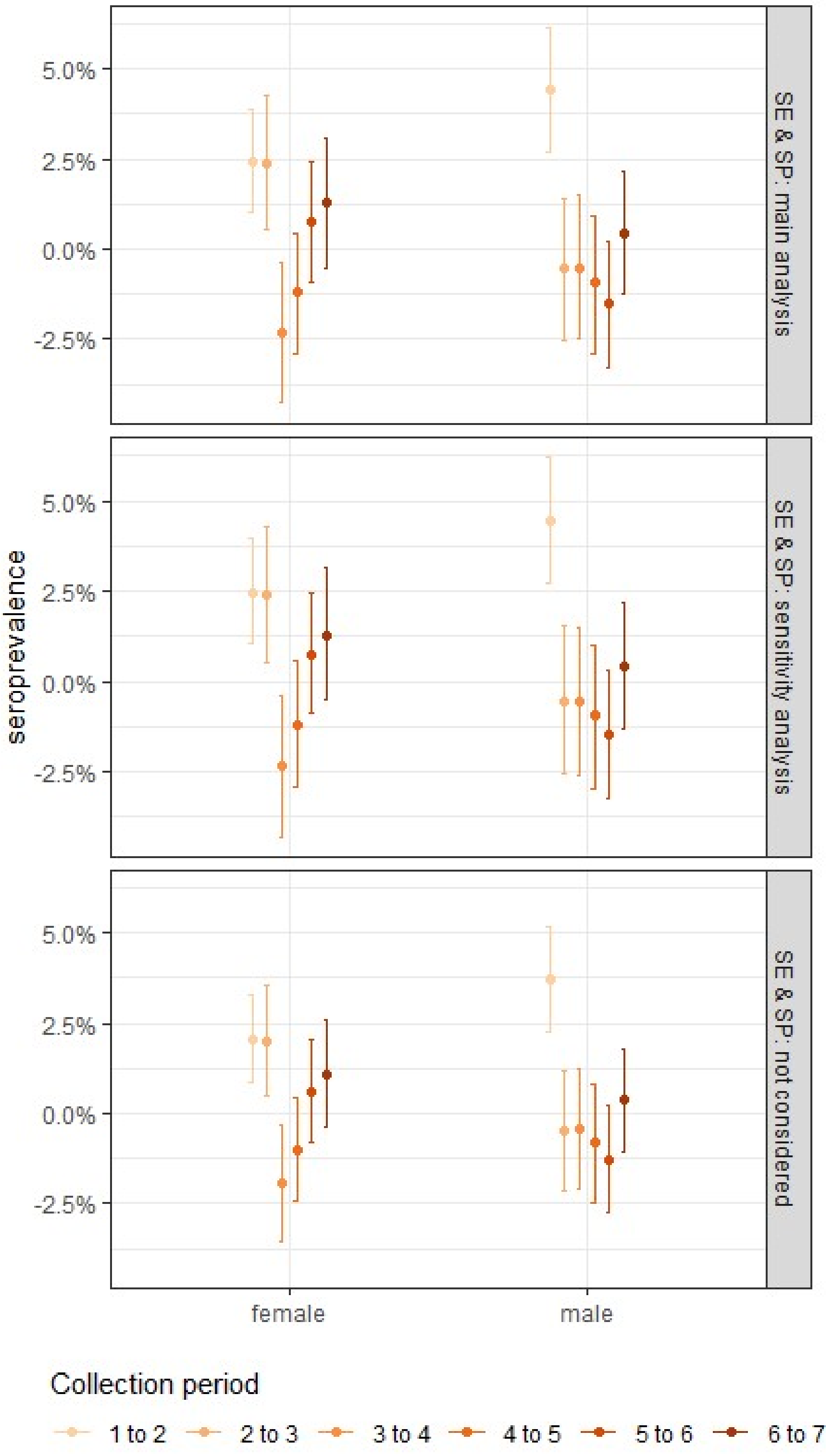

**Panel 'SE & SP: main analysis'** contains the estimates displayed in the main text (Figure 3)
**Panel 'SE & SP: sensitivity analysis'** contains the analytical sensitivity results which uses only information given for outpatient positive controls (n=90) for estimating sensitivity and specificity
**Panel 'SE & SP: not considered'** contains the estimates if sensitivity and specificity are not considered in the analysis
