## Supplementary material for "Seroprevalence of IgG antibodies against SARS coronavirus 2 in Belgium – a serial prospective cross-sectional nationwide study of residual samples (March – October 2020)": STROBE checklist

| | Item No | Recommendation | Page Number |
|---|---|---|---|
| **Title and abstract** | | | |
| | 1 | (*a*) Indicate the study's design with a commonly used term in the title or the abstract | 1 |
| | | (*b*) Provide in the abstract an informative and balanced summary of what was done and what was found | 1 |
| **Introduction** | | | |
| Background/rationale | 2 | Explain the scientific background and rationale for the investigation being reported | 2, 3 |
| Objectives | 3 | State specific objectives, including any prespecified hypotheses | 2, 3 |
| **Methods** | | | |
| Study design | 4 | Present key elements of study design early in the paper | 4, 5 |
| Setting | 5 | Describe the setting, locations, and relevant dates, including periods of recruitment, exposure, follow-up, and data collection | 4-6 |
| Participants | 6 | (*a*) *Cohort study*?Give the eligibility criteria, and the sources and methods of selection of participants. Describe methods of follow-up*Case-control study*?Give the eligibility criteria, and the sources and methods of case ascertainment and control selection. Give the rationale for the choice of cases and controls*Cross sectional study*?Give the eligibility criteria, and the sources and methods of selection of participants | 4 |
| | | (*b*) *Cohort study*?For matched studies, give matching criteria and number of exposed and unexposed*Case-control study*?For matched studies, give matching criteria and the number of controls per case | NA |
| Variables | 7 | Clearly define all outcomes, exposures, predictors, potential confounders, and effect modifiers. Give diagnostic criteria, if applicable | 6, 7 |
| Data sources/ measurement | 8* | For each variable of interest, give sources of data and details of methods of assessment (measurement). Describe comparability of assessment methods if there is more than one group | 6, 7 |
| Bias | 9 | Describe any efforts to address potential sources of bias | 6, 7 |
| Study size | 10 | Explain how the study size was arrived at | 5 |
| Quantitative variables | 11 | Explain how quantitative variables were handled in the analyses. If applicable, describe which groupings were chosen and why | 6, 7 |
| Statistical methods | 12 | (*a*) Describe all statistical methods, including those used to control for confounding | 6, 7 |
| | | (*b*) Describe any methods used to examine subgroups and interactions | 6, 7 |
| | | (*c*) Explain how missing data were addressed | NA |
| | | (*d*) *Cohort study*?If applicable, explain how loss to follow-up was addressed*Case-control study*?If applicable, explain how matching of cases and controls was addressed*Cross sectional study*?If applicable, describe analytical methods taking account of sampling strategy | NA |
| | | (*e*) Describe any sensitivity analyses | NA |
| **Results** | | | |
| Participants | 13* | (*a*) Report numbers of individuals at each stage of study?eg numbers potentially eligible, examined for eligibility, confirmed eligible, included in the study, completing follow-up, and analysed | 8 |
| | | (*b*) Give reasons for non-participation at each stage | NA |
| | | (*c*) Consider use of a flow diagram | NA |

| | | | |
|---|---|---|---|
| Descriptive data | 14* | (*a*)Give characteristics of study participants (eg demographic, clinical, social) and information on exposures and potential confounders | 8 |
| | | (*b*) Indicate number of participants with missing data for each variable of interest | NA |
| | | (*c*) *Cohort study*?Summarise follow-up time (eg average and total amount) | NA |
| Outcome data | 15* | *Cohort study*?Report numbers of outcome events or summary measures over time | NA |
| | | *Case-control study?*Report numbers in each exposure category, or summary measures of exposure | NA |
| | | *Cross sectional study?*Report numbers of outcome events or summary measures | 8 |
| Main results | 16 | (*a*) Report the numbers of individuals at each stage of the study?eg numbers potentially eligible, examined for eligibility, confirmed eligible, included in the study, completing follow-up, and analysed | 8 |
| | | (*b*) Give reasons for non-participation at each stage | NA |
| | | (*c*) Consider use of a flow diagram | NA |
| Other analyses | 17 | Report other analyses done?eg analyses of subgroups and interactions, and sensitivity analyses | 8, 9 |
| **Discussion** | | | |
| Key results | 18 | Summarise key results with reference to study objectives | 10 |
| Limitations | 19 | Discuss limitations of the study, taking into account sources of potential bias or imprecision. Discuss both direction and magnitude of any potential bias | 10-13 |
| Interpretation | 20 | Give a cautious overall interpretation of results considering objectives, limitations, multiplicity of analyses, results from similar studies, and other relevant evidence | 10-13 |
| Generalisability | 21 | Discuss the generalisability (external validity) of the study results | 10-13 |
| **Other information** | | | |
| Funding | 22 | Give the source of funding and the role of the funders for the present study and, if applicable, for the original study on which the present article is based | Title page |

Reference: https://www.bmj.com/content/335/7624/806.full?ijkey=iMuBMEW9C95tI&keytype=ref&siteid=bmjjournals
